# Characterizing the pharmacological interaction of the antimalarial combination artefenomel-piperaquine in healthy volunteers with induced blood stage *Plasmodium falciparum*

**DOI:** 10.1101/2024.02.07.24302432

**Authors:** Azrin N. Abd-Rahman, Daniel Kaschek, Anne Kümmel, Rebecca Webster, Adam J. Potter, Anand Odedra, Stephen D Woolley, Stacey Llewellyn, Lachlan Webb, Louise Marquart, Stephan Chalon, Myriam El Gaaloul, James S. McCarthy, Jörg J. Möhrle, Bridget E. Barber

## Abstract

**Background:** The combination antimalarial artefenomel-piperaquine failed to achieve target efficacy in a phase 2b study in Africa and Vietnam. We retrospectively evaluated whether characterizing the pharmacological interaction of this antimalarial combination in a volunteer infection study (VIS) would have enabled prediction of the phase 2b study results.

**Methods:** Twenty-four healthy adults enrolled over three consecutive cohorts were inoculated with *Plasmodium falciparum*-infected erythrocytes on day 0. Participants were randomized within each cohort to one of 7 dose combination groups and administered a single oral dose of artefenomel-piperaquine on day 8. Participants received definitive antimalarial treatment with artemether-lumefantrine upon parasite regrowth or on day 42±2. The General Pharmacodynamic Interaction (GPDI) model implemented in the Bliss Independence additivity criterion was developed to characterize the pharmacological interaction between artefenomel and piperaquine. Simulations based on the model were performed to predict the outcomes of the phase 2b combination study.

**Results:** For a dose of 800 mg artefenomel administered with 640 mg, 960 mg, or 1440 mg piperaquine, the simulated adequate parasitological response at day 28 (APR_28_), incorporating actual patient pharmacokinetic (PK) data from the phase 2b trial, was 69.4%, 63.9%, and 74.8%, respectively. These results closely matched the observed APR_28_ in the phase 2b trial of 67.0%, 65.5%, and 75.4% respectively.

**Conclusions:** These results indicate that VIS offer an efficient means for informing antimalarial combination trials conducted in the field, potentially expediting clinical development.

**Trial registration:** This study was registered on ClinicalTrials.gov on 31 May 2018 with registration number NCT03542149.

## BACKGROUND

Malaria remains a significant threat to global health, with an estimated 249 million cases and 608,000 deaths in 2022 [1]. Artemisinin-based combination therapies (ACTs) are the recommended first line treatment for uncomplicated *Plasmodium falciparum* malaria, although the emergence of artemisinin-resistant *P. falciparum* parasites in Southeast Asia [2], and more recently in East Africa [3–5], threatens the utility of ACTs and progress toward malaria eradication. The development of novel antimalarial therapies is required to control malaria in regions with high prevalence of artemisinin-resistance, and to prevent the spread of resistance to areas where ACTs are currently effective.

Combination therapies are the focus of antimalarial drug development, aiming to reduce the risk of selecting for resistant mutants and to target multiple stages of the parasite lifecycle including the transmissible gametocytes. Further, using a combination of two or more drugs is typically considered a prerequisite for achieving cure, given that antimalarial treatment is ideally given as a single dose or as a very short course of therapy. In the case of ACT regimens, all of which require multiple-dose treatments, the artemisinin component rapidly reduces the parasite burden, while the partner drug with a long elimination half-life is relied upon to clear residual parasites. Combination partners in approved ACTs include lumefantrine, amodiaquine, piperaquine, mefloquine, pyronaridine, and sulfadoxine-pyrimethamine [6].

Artefenomel (previously known as OZ439) is an investigational antimalarial that progressed from pre-clinical [7] to phase 1 [8,9] and phase 2 [10] clinical studies. Artefenomel is an ozonide and is thought to act in a similar manner to the artemisinins by reacting with iron within the parasite food vacuole to produce free radicals, leading to alkylation of key parasite proteins [11]. In patients with malaria, artefenomel exhibits similar antimalarial activity to artemisinins, with parasite clearance half-lives of 4.1 h to 5.6 h following single doses of 200-1200 mg [10]. However, artefenomel has a longer elimination half-life than artemisinins (approximately 50 h for artefenomel [10] compared to less than 10 h for artemisinin derivatives [12]).

Co-administration of artefenomel with piperaquine was considered a potential alternative to current ACTs, leading to testing of this combination in a phase 2b clinical trial undertaken in Africa and Vietnam [13]. Patients with uncomplicated *P. falciparum* malaria (predominantly children ≤5 years of age) were administered a single 800 mg dose of artefenomel plus piperaquine in ascending single doses (640 mg, 960 mg, or 1440 mg). Unfortunately, none of the treatment arms reached the target efficacy of >95% adequate clinical and parasitological response at day 28 (ACPR_28_), with ACPR_28_ of 70.8%, 68.4%, and 78.6% for each dose group respectively [13]. Single dose combinations that may have achieved target efficacy were retrospectively estimated via simulations using trial data. However, doses of the combination that were predicted to be required to reach this target (800 mg artefenomel plus ≥2000 mg piperaquine; 1200 mg artefenomel plus ≥960 mg piperaquine; or 1600 mg artefenomel plus ≥320 mg piperaquine) were deemed unfeasible due to safety, tolerability and practical considerations [13].

Malaria volunteer infection studies (VIS) using the induced blood-stage malaria (IBSM) model involve the inoculation of healthy, malaria naïve adults with parasites followed by administration of the test antimalarial when a predefined parasitemia threshold has been reached, and have successfully characterized the antimalarial activity of several drugs in monotherapy, including artefenomel [8] and piperaquine [14].

Pharmacokinetic/pharmacodynamic (PK/PD) modelling using data from the previously conducted artefenomel VIS involving a Caucasian volunteer population was found to accurately predict antimalarial activity in an endemic population when artefenomel monotherapy was subsequently tested in a phase 2a trial in Thailand [10]. VIS also have the potential to investigate PK/PD interactions between two or more antimalarials when administered in combination; such a study was conducted for the first time with the IBSM model to investigate artefenomel in combination with another antimalarial candidate DSM265 [15]. Data from this study were subsequently used for PK/PD simulation to predict that a single dose curative dose of 800 mg of artefenomel combined with 450 mg of DSM265 would likely achieve the target ACPR [16].

In the current study we aimed to retrospectively evaluate the pharmacological interaction between artefenomel and piperaquine in a VIS to test the hypothesis that such an approach would have predicted the outcome of the combination phase 2b study. Such an evaluation was considered important given the ethical, expense, time commitment, and safety considerations associated with a failed clinical trial of an antimalarial combination in an endemic setting.

## METHODS

### Study design and participants

This was a randomized, open label, VIS using the *P. falciparum* IBSM model. Healthy, malaria naïve, adults aged 18-55 years were eligible for inclusion (Text S1, Additional file 1). The study was conducted at Q-Pharm (Brisbane, Australia) following approval by the QIMR Berghofer Medical Research Institute Human Research Ethics Committee. All participants gave written informed consent before enrolment. This study was registered on ClinicalTrials.gov on 31 May 2018 with registration number NCT03542149.

### Procedures

The study consisted of three consecutive cohorts of 8 participants per cohort. Participants were inoculated intravenously with *P. falciparum* 3D7-infected human erythrocytes (approximately 2800 viable parasites) on day 0 and parasitemia was monitored by quantitative polymerase chain reaction (qPCR) targeting the gene encoding *P. falciparum* 18S rRNA [17]. Single oral doses of artefenomel and piperaquine phosphate were administered concurrently on day 8 in a fasted state (participants were fasted for 6 hours pre- and post-dosing). Artefenomel granules (200 mg or 400 mg) with the excipient α-tocopherol polyethylene glycol 1000 succinate (PCI Pharma Services, UK) were mixed with water to form an oral suspension, and sucrose was added prior to administration to make the oral suspension palatable. After the participant consumed the suspension, the cup was rinsed with water which was then used to facilitate swallowing the piperaquine tablets (160 mg per tablet; PCI Pharma Services, UK). Participants were confined to the clinic for 72 h post-dosing and returned as outpatients for follow up visits until the end of study visit on day 45±2. Participants received a standard curative course of artemether-lumefantrine (Riamet^®^; Novartis Pharmaceuticals Pty Ltd, Australia) upon parasite regrowth, or on day 42±2 if parasite regrowth had not occurred.

Blood samples were collected pre-dose and at the following time-points after dosing to determine artefenomel and piperaquine plasma concentration: 0.5, 1, 2, 3, 4, 5, 6, 8, 12, 16, 24, 48, 72, 96, 168, 240, 336, 504, 672, and 840 hours. Drug concentrations were determined using liquid chromatography tandem mass spectrometry as described previously for artefenomel [9] and piperaquine [18]. To monitor total parasitemia by 18S qPCR, blood samples were collected before inoculation on day 0, day 4, twice daily on days 5-7, pre-dosing and 2, 4, 8, 12, 16, 20, 24, 30, 36, 42 hours post-dosing on day 8 and 9, twice daily on days 10-13, and every 1 to 3 days until the end of study (day 45±2).

### Sample size, randomization, and dose selection

The intended sample size (24 participants) was not based on a formal power calculation but, based on previous studies [15], was considered adequate to assess the primary objective of the study. More than one dose level was tested within each cohort to optimally characterize the PK/PD relationship of the combination. Participants were randomized within each cohort to a dose group on the day of dosing. Cohort 1 consisted of 4 dose groups while cohort 2 and cohort 3 consisted of 2 dose groups. Randomization was balanced over the dose groups within each cohort. The randomization schedule was generated using Stata version 15 (StataCorp, College Station, TX, USA). No blinding was performed.

The doses of artefenomel and piperaquine were chosen to facilitate modelling to estimate the PK/PD relationship. For cohort 1, doses were based on the PK/PD relationships observed in IBSM monotherapy studies of artefenomel [8] and piperaquine [14] and the potential PD interaction effects of the combination based on the phase 2b combination trial [13]. The doses for cohorts 2 and 3 were decided based on a preliminary PK/PD model which utilized data from the preceding cohort and from the artefenomel and piperaquine monotherapy VIS. Different dose combinations were evaluated using popED [19] assuming known PK parameters. Estimation errors for the PD parameters and the D-optimality criterion were considered [19]. Promising doses based on this evaluation for the next cohort were simulated and doses selected following discussion between the modelers and study investigators.

### Pharmacokinetic/pharmacodynamic modelling

The VIS PK/PD model was built in a stepwise manner, using data from the current study as well as data from the artefenomel [8] and piperaquine [14] monotherapy VIS (Table S1, Additional file 2). A population PK model was firstly developed to obtain individual PK parameter estimates which adequately described the observed individual PK profiles. Drug-drug interaction on the PK parameters was tested as inhibition of the clearance of one drug by the concentrations of the other drug. The PK/PD model was then built using the individual PK parameter estimates as regressors to evaluate the relationship between artefenomel and piperaquine plasma concentration and parasite killing. A PD model for each compound alone was built independently to estimate the single-drug effect parameters. The combined effects of artefenomel and piperaquine were evaluated using the General Pharmacodynamic Interaction model (GPDI model [20]) implemented in the Bliss Independence additivity criterion. Model evaluation and selection was guided by visual inspection of goodness of fit plots, of individual PK and PD profiles, plausibility and precision of parameter estimates, and fit statistics such as Bayesian information criterion.

### Simulations of clinical success in patients

To assess the translatability of the VIS PK/PD data, the adequate parasitological response at day 28 (APR_28_) was predicted based on a PK/PD model and compared with corresponding observations from the Phase 2b trial of the artefenomel + piperaquine combination [13]. APR_28_ was defined as a solely parasitemia based simplification of the ACPR_28_ clinical success criterion [21], in the absence of body temperature data relevant for the evaluation of ACPR_28_ (Text S2, Additional file 1).

Two separate simulations were performed using either the PK model derived from VIS data, or actual PK data from patients in the Phase 2b trial (in which case the estimated individual patient PK parameters were fixed as regressors). For both simulations, the PD parameters for artefenomel and piperaquine were sampled based on the respective VIS PK/PD monotherapy models, and the PD interaction parameters were sampled based on the VIS GPDI model. The sampling of the VIS PD models and interaction parameters (and VIS PK model where appropriate) was done in two steps. First, 250 sets of reference population parameter values were sampled from the parameter uncertainty distribution. Then, for each such set, a population of individual parameter sets was sampled from the inter-individual variability distribution. The number of patients in each simulated population matched the population size of the phase 2b trial (Table S2, Additional file 2). Simulations used the actual baseline parasitaemia values recorded for patients in the phase 2b trial, and patient body weight was accounted for in simulations using the VIS PK model (Table S2, Additional file 2). Parasites were assumed to grow exponentially in patients at a growth rate of 0.048 hours^−1^ (equivalent to a multiplication rate of ∼ 10-fold per 48-hour asexual cycle) [22,23].

The APR_28_ was determined for the observed and simulated parasitemia time courses. For the observed parasitemia, the proportion of patients with APR_28_ status was reported and the 95% CI was estimated using the Clopper-Pearson method. For the simulated parasitemia, the median APR_28_ proportion over the 250 populations was reported and the 0.025^th^ and 0.975^th^ quantiles were used as an approximation of the 95% CI.

All data processing, analysis, model setup and modelling analysis, including goodness-of-fit plots, was performed in R 3.5.1 using the IQRtools package 1.0.0 (https://iqrtools.intiquan.com). Nonlinear mixed effects (NLME) modelling was performed with Monolix 2018R2 (http://lixoft.com) using Stochastic Approximation Expectation Maximization (SAEM) for parameter estimation.

### Non-compartmental pharmacokinetic analysis

Non-compartmental PK parameters calculated were: maximum observed concentration (C_max_), time to reach C_max_ (t_max_), area under the concentration-time curve (AUC) from time 0 (dosing) to the last sampling time at which the concentration is ≥the lower limit of quantification (LLOQ) (AUC_0-last_), AUC from time 0 extrapolated to infinity (AUC_0-inf_), apparent elimination half-life (t_½_), apparent total body clearance (CL/F), apparent total volume of distribution (Vz/F), elapsed time from dosing at which drug concentration was first quantifiable (t_lag_), and the apparent terminal elimination rate constant (λ_inf_). Non-compartmental pharmacokinetic analysis was performed in R version 3.5.1 (https://R-project.org) using the IQR tools package 1.0.0.

### Parasite clearance analysis

The parasite reduction ratio (PRR) and corresponding parasite clearance half-life (PCt_1/2_) were estimated, with the former expressed as the ratio of the parasite density decrease over 48 hours following dosing (PRR_48_). The PRR_48_ and parasite clearance half-life were estimated using the slope of the optimal fit for the log-linear relationship of the parasitemia decay as described previously [24]. Briefly, the decay rate (slope coefficient from the log-linear decay regression) for each participant was calculated initially, then the weighted average slope estimate and corresponding standard error was calculated using an inverse-variance method, which was used to estimate the dose specific PRR_48_ and 95% CI. The PCt_1/2_ is a transformation of the slope coefficient into a time period. Parasite clearance analyses were performed in R version 3.5.1. The percentage of participants with parasite regrowth following dosing was also calculated.

### Safety and tolerability analysis

The incidence, severity and relationship to artefenomel + piperaquine administration of adverse events (AEs) was monitored. The period of observation for collection of AEs extended from the time of inoculation with the malaria challenge agent up to the end of the study. AE severity was assessed in accordance with the Common Terminology Criteria for Adverse Events [25] (mild=grade 1; moderate=grade 2; severe=grade 3; life-threatening consequences=grade 4; death related to AE=grade 5). In addition, an AE was classified as a serious adverse event (SAE) if it met one of the following criteria: resulted in death, was life-threatening, required inpatient hospitalization, resulted in persistent or significant disability, was a congenital anomaly, was considered medically important, constitutes a possible Hy’s Law case. The investigator assessed if AEs were related to artefenomel + piperaquine and/or to the malaria challenge (unrelated, unlikely, possible, probable). Safety assessments included clinical laboratory parameters (hematology, biochemistry, and urinalysis), vital signs (body temperature, blood pressure, heart rate, respiratory rate), physical examination, and 12-lead electrocardiographs (ECGs).

## RESULTS

### Participant disposition

The study was conducted between April 2018 and April 2019. A total of 24 healthy participants in three consecutive cohorts of eight participants were enrolled and inoculated with *P. falciparum*-infected erythrocytes on day 0 (Figure 1). Following inoculation, participants were randomized within each cohort (non-blinded) to a dose group. Dose combinations tested in cohort 1 were pre-defined in the study protocol while dose combinations tested in cohorts 2 and 3 were decided following preliminary PK/PD data assessment from the previous cohort. All participants received the allocated single oral dose of artefenomel and piperaquine on day 8. Two groups received the same dose (cohort 1D and cohort 3A; 400 mg artefenomel and 640 mg piperaquine) and were combined for data analysis. Most participants were male (18/24, 75.0%) and self-selected their race as white (19/24, 79.2%); the mean age of participants was 25.0 years (Table 1).

**Figure 1.**
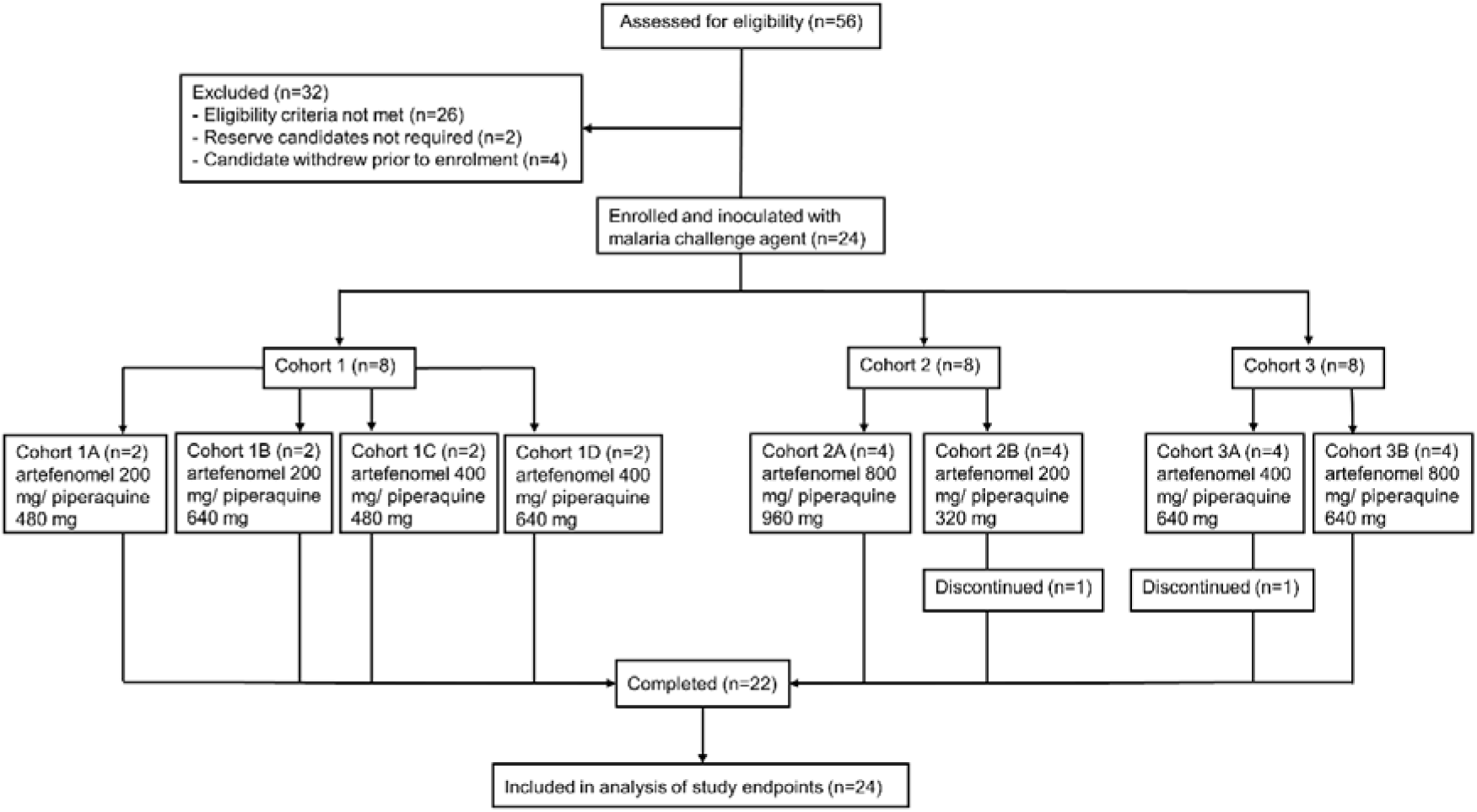
Trial profile. Enrolled participants were randomized within each cohort to a dose group on the day of dosing with artefenomel + piperaquine combination treatment (8 days following challenge with blood-stage *P. falciparum*). Two participants discontinued voluntarily prior to the end of study; available data from both participants were included in the analysis of study endpoints.

**Table 1.** Demographic profile of participants.

| Parameter |  | Artefenomel<br>200 mg +<br>piperaquine<br>480 mg<br>[N=2] | Artefenomel<br>200 mg +<br>piperaquine<br>640 mg<br>[N=2] | Artefenomel<br>400 mg +<br>piperaquine<br>480 mg<br>[N=2] | Artefenomel<br>400 mg +<br>piperaquine<br>640 mg<br>[N=6] | Artefenomel<br>800 mg +<br>piperaquine<br>960 mg<br>[N=4] | Artefenomel<br>200 mg +<br>piperaquine<br>320 mg<br>[N=4] | Artefenomel<br>800 mg +<br>piperaquine<br>640 mg<br>[N=4] | Total<br>[N=24] |
| --- | --- | --- | --- | --- | --- | --- | --- | --- | --- |
| Age [years] | Mean (SD) | 36.5 (7.8) | 23.0 (1.4) | 40.5 (17.7) | 23.5 (4.1) | 21.3 (2.2) | 23.8 (3.6) | 20.0 (1.4) | 25.0 (7.9) |
| Sex [n (%)] | Male | 2 (100) | 2 (100) | 1 (50.0) | 6 (100) | 3 (75.0) | 2 (50.0) | 2 (50.0) | 18 (75.0) |
|  | Female | 0 | 0 | 1 (50.0) | 0 | 1 (25.0) | 2 (50.0) | 2 (50.0) | 6 (25.0) |
| Race [n (%)] | White | 1 (50.0) | 0 | 2 (100) | 5 (83.3) | 3 (75.0) | 4 (100) | 4 (100) | 19 (79.2) |
|  | Asian | 1 (50.0) | 2 (100) | 0 | 1 (16.7) | 0 | 0 | 0 | 4 (16.7) |
|  | Multiple races | 0 | 0 | 0 | 0 | 1 (25.0) | 0 | 0 | 1 (4.2) |
| BMI [kg/m<sup>2</sup>] | Mean (SD) | 25.0 (3.4) | 22.5 (2.4) | 22.1 (2.2) | 23.2 (2.4) | 22.0 (2.7) | 25.1 (1.1) | 23.1 (3.1) | 23.3 (2.4) |
| Height (cm) | Mean (SD) | 179.5 (16.3) | 172.5 (0.7) | 174.0 (4.2) | 179.5 (8.7) | 178.5 (7.9) | 175.3 (6.3) | 174.0 (5.5) | 176.7 (7.3) |
| Weight (kg) | Mean (SD) | 81.9 (25.4) | 66.9 (6.7) | 67.0 (9.9) | 74.5 (6.5) | 70.0 (9.6) | 77.0 (4.6) | 70.1 (10.9) | 72.8 (9.7) |

BMI: body mass index; SD: standard deviation.

### Parasitemia and drug exposure

The progression of parasitemia following inoculation with blood-stage *P. falciparum* was consistent between participants, with an expected increase in parasitemia up to artefenomel + piperaquine administration on day 8 (Figure 2). Dose-related increases in artefenomel and piperaquine exposure were observed across the dose range tested (Figure 3A/B). Maximum plasma concentrations occurred 2-3 h after dosing for artefenomel and 3-5 h after dosing for piperaquine (Table S4, Additional file 2). An initial rapid fall in parasitemia occurred in all participants following dosing (Figure 2), with a trend of increased rate of parasite clearance with increased dose of the combination (Table 2 and Table S5, Additional file 2). Parasite regrowth occurred after dosing with artefenomel 200 mg + piperaquine 320 mg (4/4 participants; 5 days post-dose), artefenomel 200 mg + piperaquine 480 mg (2/2 participants; 4 and 14 days post-dose), and artefenomel 400 mg + piperaquine 480 mg (1/2 participants; 14 days post-dose). Parasite regrowth was not observed in any of the other dose groups up to 35±2 days post-dosing when definitive antimalarial treatment with artemether-lumefantrine was initiated. One participant in the artefenomel 400 mg + piperaquine 640 mg group voluntarily withdrew from the study 7 days post-dosing requiring early artemether-lumefantrine treatment. One participant in the artefenomel 200 mg + piperaquine 320 mg group voluntarily withdrew from the study 3 days after artemether-lumefantrine had been administered for parasite regrowth. Available data from both participants who withdrew prior to the end of study were included in the PK/PD analysis. All participants were confirmed to be aparasitemic by the end of the study.

**Figure 2.**
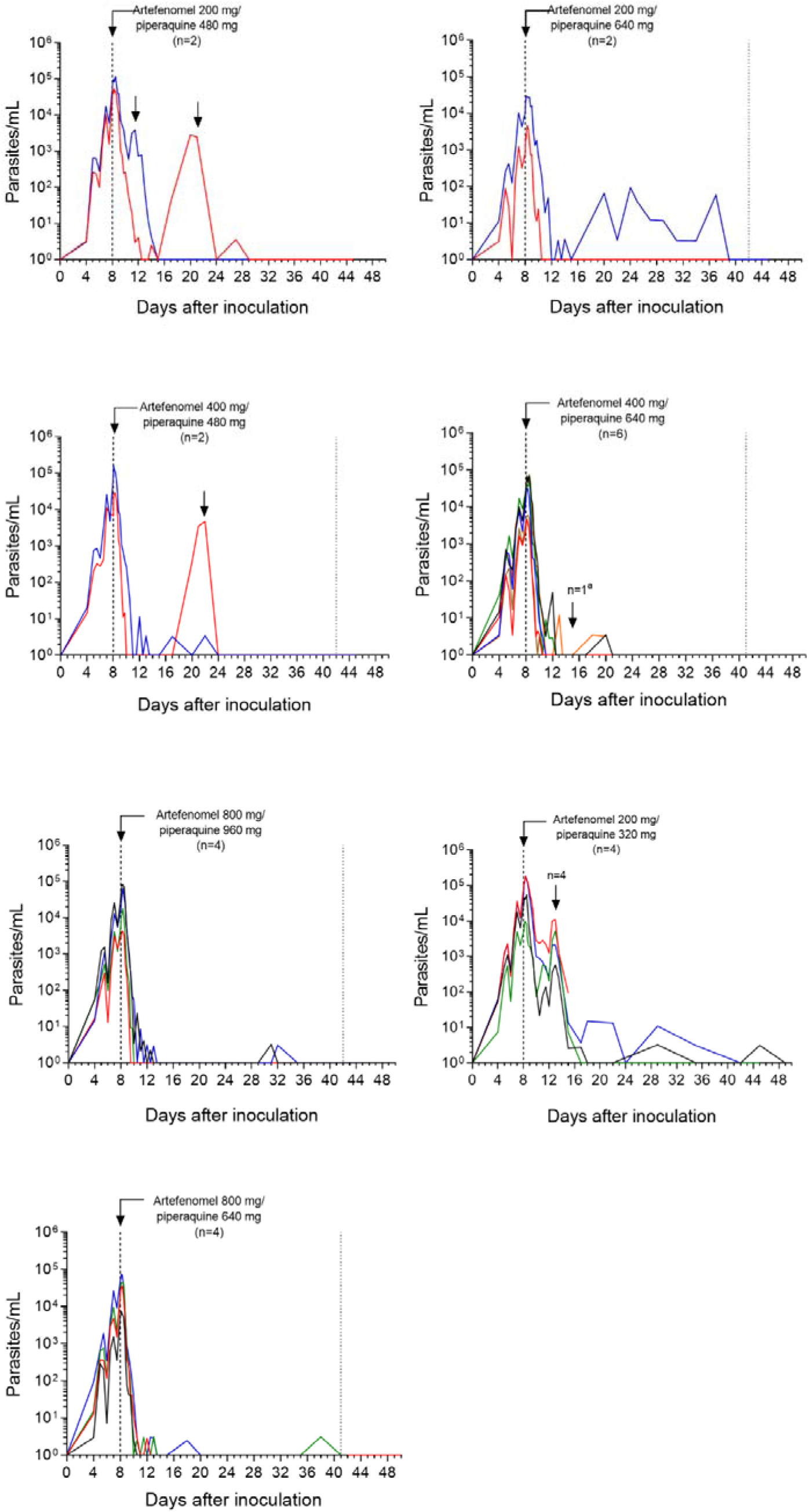
Individual participant parasitemia-time profiles. Participants were inoculated intravenously with *P. falciparum*-infected erythrocytes and were administered a single oral dose of artefenomel and piperaquine in combination after 8 days (indicated by the vertical dashed line). Parasitemia was monitored using qPCR targeting the gene encoding *P. falciparum*18S rRNA. Artemether-lumefantrine was administered in response to parasite regrowth (indicated by the vertical arrows) or 35±3 days after artfenomel + piperaquine dosing if parasite regrowth was not observed (indicated by the vertical dotted line). For the purpose of graphing on a log_10_ logarithmic scale, time points at which parasitemia could not be detected were substituted with a value of 1 parasite/mL. ^a^One participant received early artemether-lumefantrine treatment due to voluntary withdrawal from the study.

**Figure 3.**
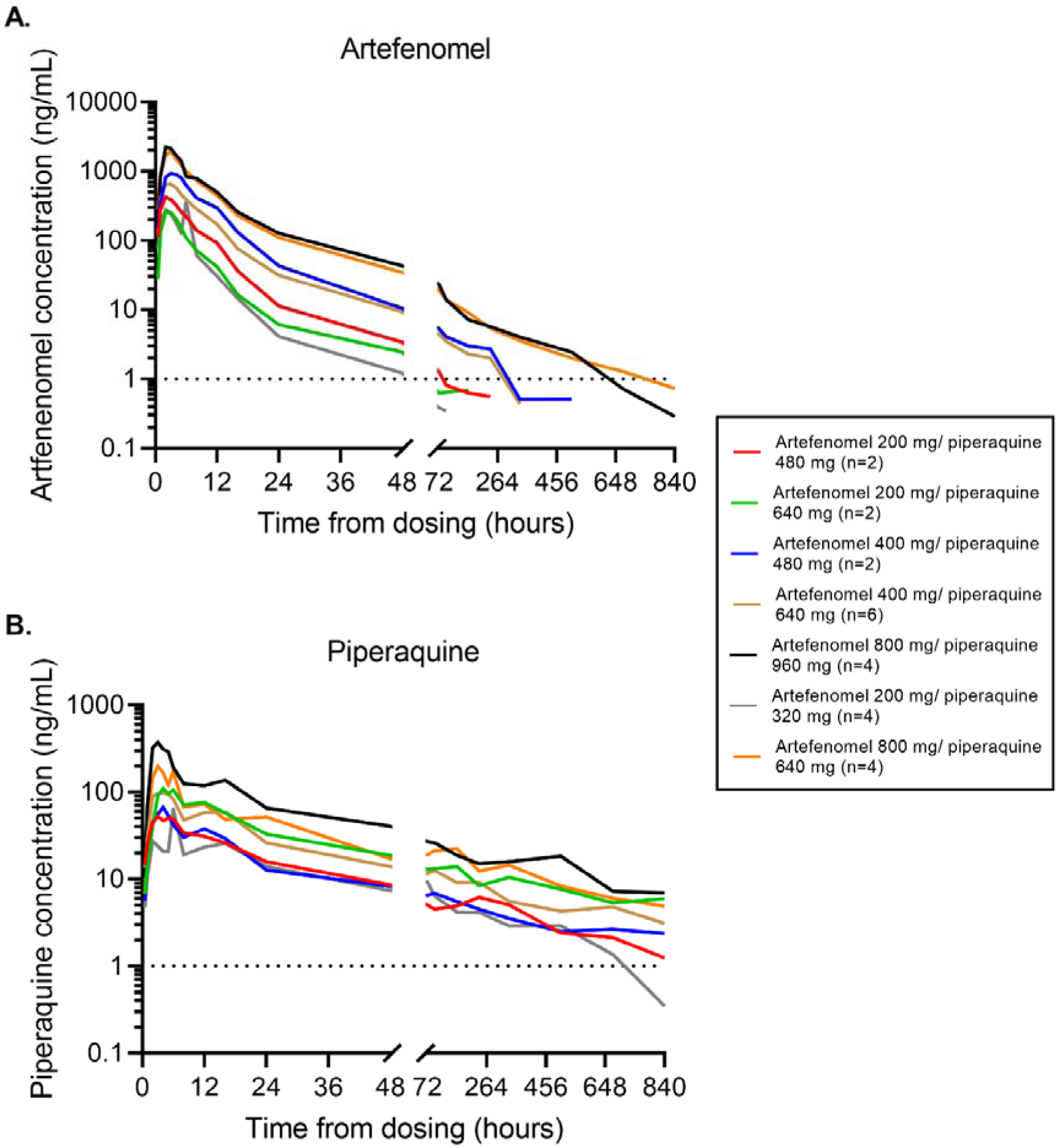
Artefenomel and piperaquine plasma concentration-time profiles. Plots represent the arithmetic mean of the artefenomel (A) and piperaquine (B) plasma concentration of each dose group over the study. Plasma concentrations were measured using liquid chromatography with tandem mass spectrometry. The horizontal dotted lines indicate the lower limit of quantification (1 ng/mL).

**Table 2.** Parasite clearance kinetics and incidence of parasite regrowth.

| Treatment Group | Log<sub>10</sub> PRR<sub>48</sub> (95% CI) | PC<sub>t1/2</sub> [hours] (95% CI) | Parasite regrowth [n (%)] |
| --- | --- | --- | --- |
| Artefenomel 200 mg + piperaquine 480 mg [N=2] | 2.18 (2.00 - 2.36) | 6.63 (6.13 - 7.22) | 2 (100) |
| Artefenomel 200 mg + piperaquine 640 mg [N=2] | 2.34 (2.08 - 2.60) | 6.18 (5.57 - 6.93) | 0 |
| Artefenomel 400 mg + piperaquine 480 mg [N=2] | 3.58 (3.29 - 3.86) | 4.04 (3.74 - 4.39) | 1 (50.0) |
| Artefenomel 800 mg +/- piperaquine 960 mg [N=4] | 3.98 (3.74 - 4.23) | 3.63 (3.41 - 3.87) | 0 |
| Artefenomel 200 mg +/- piperaquine 320 mg [N=4] | 1.89 (1.75 - 2.04) | 7.63 (7.10 - 8.25) | 4 (100) |
| Artefenomel 800 mg + piperaquine 640 mg [N=4] | 3.56 (3.36 - 3.76) | 4.06 (3.84 - 4.30) | 0 |
| Artefenomel 400 mg + piperaquine 640 mg [N=6] | 3.70 (3.52 - 3.88) | 3.90 (3.72 - 4.10) | 0 |

PRR<sub>48</sub>: parasite reduction ratio over a 48 h period; PC<sub>t1/2</sub>: parasite clearance half-life.

### Pharmacokinetic/pharmacodynamic modelling

The VIS PK/PD model was built using data from two prior VIS, in which artefenomel [8] and piperaquine [14] were administered as monotherapy, as well as data obtained in the current study. Population PK modelling using VIS data indicated that the PK profiles of artefenomel and piperaquine were appropriately described by a two-compartmental distribution model with linear elimination and first order absorption (parameter estimates presented in Table S6, Additional file 2; visual predictive checks presented in Figures S1-S4, Additional file 3).

Models including inhibition of piperaquine clearance by artefenomel, inhibition of artefenomel clearance by piperaquine, or inhibition of clearance of both drugs by the other were tested. As none of the inhibition terms improved the model fit as measured by Bayesian information criterion, no PK drug-drug interaction was included in the model. The PD of artefenomel and piperaquine were described by sigmoidal (E_max_) concentration-killing relationships, with parameters estimated based on the respective monotherapy VIS data (Table S7, Additional file 2). The concentrations with half-maximal killing effect (EC_50_) were estimated to be 2.89 ng/mL for artefenomel and 6.26 ng/mL for piperaquine. The maximum killing rate (E_max_) for each drug were comparable (0.194/h for artefenomel and 0.262/h for piperaquine). A GPDI model, quantifying the mutual modulation of EC_50_ and E_max_, described the parasitemia time courses for combination treatment (Figure S5, Additional file 3). In this model, the estimated interaction parameters were negative (Table S7, Additional file 2), suggesting synergy of both potency and efficacy for the two compounds in combination. The interaction parameters of the GPDI model were estimated with small uncertainty given the relative standard error was less than 30% (Table S7, Additional file 2).

### Simulation of clinical success rates in patients

The probability of adequate parasitological response at day 28 (APR_28_) was simulated for patients with acute uncomplicated *P. falciparum* malaria enrolled in the phase 2b trial of artefenomel + piperaquine combination therapy using a PK/PD model incorporating the PD interaction parameters estimated from the current VIS and individual drug PD parameters estimated from VIS monotherapy data. These PD parameters were combined with either the PK data derived from the VIS, or in a separate simulation, with actual PK data derived from the patients in the Phase 2b trial. Simulations were compared with the observed clinical success rates in the phase 2b trial to determine the predictive performance of the model. Simulations incorporating the VIS PK data highly overestimated the observed clinical success rates, with APR_28_ rates of 100% for each of the three dose combination groups (Table 3). This result was consistent with the fact that the plasma levels achieved for both artefenomel and piperaquine were higher in VIS participants than those achieved in patients in the phase 2b trial for equivalent dose combinations (Figures S6 and S7, Additional file 3). Simulations incorporating observed PK data from patients in the Phase 2b trial predicted the probability of APR_28_ across the three treatment groups with a high degree of accuracy (Table 3). The observed APR_28_ rate in the phase 2b trial was 67.0% for artefenomel 800 mg + piperaquine 640 mg, 65.5% for artefenomel 800 mg + piperaquine 960 mg, and 75.4% for artefenomel 800 mg + piperaquine 1440 mg. The simulated APR_28_ rates were 69.4%, 63.9%, and 74.8%, respectively.

**Table 3.** Probability of APR_28_ based on patient data from a phase 2b combination trial compared to PK/PD model simulations.

|  | Clinical trial | PK/PD model (VIS PK) | PK/PD model (Patient PK) |
| --- | --- | --- | --- |
| Artefenomel 800 mg + piperaquine 640 mg | 67.0 (57.2 – 75.8) | 100 (98.1 – 100)* | 69.4 (65.4 – 74.1) |
| Artefenomel 800 mg + piperaquine 960 mg | 65.5 (56.3 – 74.0) | 100 (99.2 – 100)* | 63.9 (60.5 – 68.9) |
| Artefenomel 800 mg + piperaquine 1440 mg | 75.4 (66.6 – 82.9) | 100 (99.2 – 100)* | 74.8 (74.8 – 80.7) |

Estimated probability is denoted in % with 95% confidence interval in parentheses. For clinical trial data, the estimated probability of event occurrence and the 95% binomial confidence intervals are shown. For the PK/PD simulations, the mean probability of event occurrence 0.025th and 0.975th quantile from 250 simulated trials are shown. The results of each PK/PD model simulation were compared to clinical trial data using a two-proportions z-test with the Benjamini-Hochberg procedure (\*indicates p-value <0.05). APR<sub>28</sub>: adequate parasitological response on day 28.

### Safety and tolerability

A total of 101 AEs were reported during the study, with 21/24 participants (87.5%) experiencing at least one AE (Table 4 and Table S3, Additional file 2). All AEs were mild or moderate in severity; none were graded as severe or met the criteria for a SAE. There was no obvious relationship between the dose of artefenomel + piperaquine and the incidence of AEs. The majority of AEs were considered related to malaria (67/101 AEs). There were 7 AEs considered related to artefenomel + piperaquine dosing, experienced by 5 participants over two dose groups (artefenomel 800 mg + piperaquine 960 mg and artefenomel 400 mg + piperaquine 480 mg). These AEs were 5 events of mild nausea and one event each of mild headache and mild QT prolongation on ECG. The case of QT prolongation occurred in a participant dosed with 800 mg artefenomel and 960 mg piperaquine; the participant had a pre-dose QT interval (heart rate corrected using Fridericia’s formula; QTcF) of 422 ms which increased to 450 ms and 460 ms at 4 and 6 hours post-dosing respectively, before normalizing at 8 hours post-dosing (425 msec).

**Table 4.**
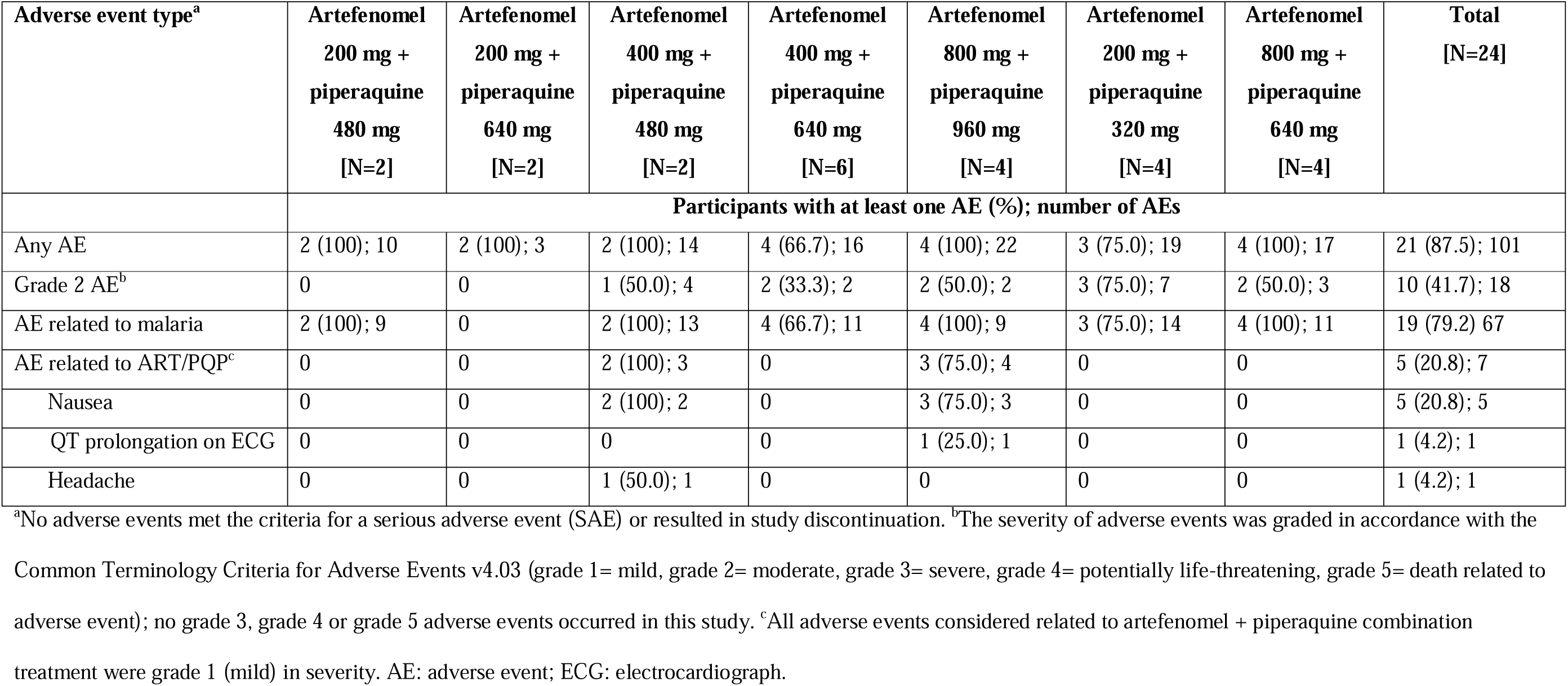
Summary of adverse events.

| Adverse event type<sup>a</sup> | Artefenomel<br>200 mg +<br>piperaquine<br>480 mg<br>[N=2] | Artefenomel<br>200 mg +<br>piperaquine<br>640 mg<br>[N=2] | Artefenomel<br>400 mg +<br>piperaquine<br>480 mg<br>[N=2] | Artefenomel<br>400 mg +<br>piperaquine<br>640 mg<br>[N=6] | Artefenomel<br>800 mg +<br>piperaquine<br>960 mg<br>[N=4] | Artefenomel<br>200 mg +<br>piperaquine<br>320 mg<br>[N=4] | Artefenomel<br>800 mg +<br>piperaquine<br>640 mg<br>[N=4] | Total<br>[N=24] |
| --- | --- | --- | --- | --- | --- | --- | --- | --- |
|  | Participants with at least one AE (%); number of AEs |  |  |  |  |  |  |  |
| Any AE | 2 (100); 10 | 2 (100); 3 | 2 (100); 14 | 4 (66.7); 16 | 4 (100); 22 | 3 (75.0); 19 | 4 (100); 17 | 21 (87.5); 101 |
| Grade 2 AE<sup>b</sup> | 0 | 0 | 1 (50.0); 4 | 2 (33.3); 2 | 2 (50.0); 2 | 3 (75.0); 7 | 2 (50.0); 3 | 10 (41.7); 18 |
| AE related to malaria | 2 (100); 9 | 0 | 2 (100); 13 | 4 (66.7); 11 | 4 (100); 9 | 3 (75.0); 14 | 4 (100); 11 | 19 (79.2) 67 |
| AE related to ART/PQP<sup>c</sup> | 0 | 0 | 2 (100); 3 | 0 | 3 (75.0); 4 | 0 | 0 | 5 (20.8); 7 |
| Nausea | 0 | 0 | 2 (100); 2 | 0 | 3 (75.0); 3 | 0 | 0 | 5 (20.8); 5 |
| QT prolongation on ECG | 0 | 0 | 0 | 0 | 1 (25.0); 1 | 0 | 0 | 1 (4.2); 1 |
| Headache | 0 | 0 | 1 (50.0); 1 | 0 | 0 | 0 | 0 | 1 (4.2); 1 |

<sup>a</sup>No adverse events met the criteria for a serious adverse event (SAE) or resulted in study discontinuation. <sup>b</sup>The severity of adverse events was graded in accordance with the Common Terminology Criteria for Adverse Events v4.03 (grade 1= mild, grade 2= moderate, grade 3= severe, grade 4= potentially life-threatening, grade 5= death related to adverse event); no grade 3, grade 4 or grade 5 adverse events occurred in this study. <sup>c</sup>All adverse events considered related to artefenomel + piperaquine combination treatment were grade 1 (mild) in severity. AE: adverse event; ECG: electrocardiograph.

Decreased white blood cell counts were observed in several participants; these were mild to moderate in severity and typically occurred within four days following dosing. Decreases in lymphocyte counts occurred in four participants (lowest nadir of the four: 0.55×10^9^/L; normal range 1.0-4.0×10^9^/L), decreases in neutrophil counts in two participants (lowest nadir of the two: 1.22×10^9^/L; normal range 1.5-8.0×10^9^/L), with a composite decrease in leukocyte count in one participant (2.9×10^9^/L; normal range 3.5-12.0×10^9^/L). Additionally, a mild decrease in hemoglobin from baseline (day 0, malaria challenge) was recorded for 4 participants (fractional fall 15-18% at nadir). These clinical laboratory abnormalities were all transient and considered to be probably or possibly related to malaria, and unrelated to combination dosing. No clinically significant elevations in liver function enzymes were recorded.

## DISCUSSION AND CONCLUSIONS

The purpose of this study was to retrospectively evaluate whether characterizing the pharmacological interaction of artefenomel and piperaquine in a small number of healthy, malaria naive adult volunteers using the *P. falciparum* IBSM model could have predicted the results of a large phase 2b combination study [13]. The phase 2b study was conducted in 7 countries over two continents (Africa and Asia). It enrolled a total of 448 patients (the majority of whom were children under the age of 5 years), with patient recruitment and follow-up occurring over a 12-month period. The negative outcome of the trial was a significant disappointment given the implications with respect to the development of an alternative treatment to ACTs and overall progress towards the goal of malaria eradication. If data from the artefenomel + piperaquine combination VIS could have been used to predict the outcome of the phase 2b study, it could be useful for informing future combination development programs, and thus optimizing the use of resources.

The pharmacological interaction between artefenomel and piperaquine was characterized using an adaptive study design to test multiple dose combinations, with dose selection informed by preliminary analyses of preceding dose groups. PK/PD modelling activities were also dependent on having the PK and PD data on each compound administered alone, obtained from prior *P. falciparum* IBSM VIS [8,14]. Consistent with previous *in vitro* drug-drug interaction assessment and the population PK model built from the phase 2b combination trial [13], no PK drug-drug interaction between artefenomel and piperaquine was identified using population PK modelling in the current study. A PK/PD interaction model (GPDI model) described the parasitemia time courses following combination treatment and indicated synergistic activity between the two drugs. This interaction model was combined with PD models of artefenomel and piperaquine generated from previous monotherapy VIS data for simulations to predict clinical success rates at day 28 for the completed phase 2b combination trial.

Notably, the accuracy of the prediction was found to be highly dependent upon the source of the PK data used in the simulations. When the PK model built from VIS data was extrapolated to the patient population, the simulations greatly overestimated clinical success compared to the observed results in the phase 2b trial. This was due to the fact that both artefenomel and piperaquine plasma exposures were higher in VIS participants compared with the patient population. When the PK model incorporated actual PK data from patients in the phase 2b trial rather than PK data from the VIS participants, the simulations were highly accurate in predicting the observed clinical success rates. The fact that there were differences in PK between the healthy adult participants (predominantly Caucasian) in the VIS and the African and Asian patients with uncomplicated *P. falciparum* malaria (predominantly children ≤5 years of age) is not unexpected. Although body weight was accounted for when using the VIS PK model for simulations in patients, other physiological factors are likely responsible for the differences in observed PK profiles.

This study has demonstrated that characterizing the pharmacological interaction of artefenomel + piperaquine in a VIS would have predicted the outcome of the selected dose combinations in the phase 2b study, provided that PK data from the target patient population were available. Dose selection in the phase 2b artefenomel + piperaquine combination was primarily designed to achieve the maximum exposure that would be well tolerated, with minimization of a potential prolongation of the QT interval by high plasma levels of piperaquine a key consideration [13,26]. The value of assessing the pharmacological interaction between antimalarial combinations during *in vitro* parasite culture, and in the humanized mouse model of malaria, to inform clinical decision making around drug and dose selection has recently been established [27,28]. However, the translatability of these models is likely to be limited by the complex PK/PD interactions that may occur in humans. Therefore, performing a VIS to characterize the PD interaction of a promising new antimalarial combination, in conjunction with a PK bridging study in the target patient population, may be a worthy strategy to optimize resources and maximize successful outcomes of subsequent phase 2 and 3 trials.

## Data Availability

The datasets used and analysed during the current study are available from the corresponding author on reasonable request.

## LIST OF ABBREVIATIONS

ACT: artemisinin-based combination therapy
ACPR_28_: adequate clinical and parasitological response at day 28
AEs: adverse events
APR_28_: adequate parasitological response at day 28
EC_50_: concentration with half-maximal killing effect
ECG: electrocardiograph
E_max_: maximum killing rate
GPDI: general pharmacodynamic interaction
IBSM: induced blood-stage malaria
LLOQ: lower limit of quantification
PC_t1/2_: parasite clearance half-life
PD: pharmacodynamic
PK: pharmacokinetic
PRR_48_: parasite reduction ratio over 48 hours
qPCR: quantitative polymerase chain reaction
SAE: serious adverse event
VIS: volunteer infection study.

## DECLARATIONS

### Ethics approval and consent to participate

The study was approved by the QIMR Berghofer Medical Research Institute Human Research Ethics Committee (reference number: P2370). All participants gave written informed consent before enrolment.

### Consent for publication

Not applicable.

### Authors’ contributions

ANA, DK, and AK performed the pharmacokinetic/pharmacodynamic modeling. AO, SW, and BEB performed the clinical components of the study. SL, LW and LM performed the analysis of parasite clearance. SC was responsible for medical oversight. MEG, JSM and JJM were responsible for study conceptualization and study design. RW performed project management activities. AJP led manuscript development. All authors read and approved the final manuscript.

## Acknowledgements

We would like to thank the volunteers who participated in the study; the clinical study team at Q-Pharm who conducted the study; staff at the Queensland Paediatric Infectious Diseases laboratory for qPCR analysis; Michael Marx from ICON Clinical Research for serving as medical monitor; Nathalie Gobeau from Medicines for Malaria Venture for contributing expertise in PK/PD modelling activities, and from the QIMR Berghofer Medical Research Institute we thank Helen Jennings, Jeremy Gower, Hayley Mitchell, and Rebecca Pawliw for preparing the malaria challenge agent.

## ADDITIONAL FILES

- **Additional file 1.docx**

- **Text S1.** Participant eligibility criteria.
- **Text S2.** Definition of adequate parasitological response on day 28 (APR_28_).
- **Additional file 2.docx**

- **Table S1.** Volunteer infection study data used for pharmacokinetic/pharmacodynamic modelling of artefenomel-piperaquine combination.
- **Table S2.** Phase 2b study data used in simulations to predict APR_28_ in patients.
- **Table S3.** Adverse events by system organ class and preferred term.
- **Table S4.** Plasma artefenomel and piperaquine non-compartmental pharmacokinetic parameters.
- **Table S5.** Individual participant parasite clearance parameters.
- **Table S6.** Parameter estimates for the final pharmacokinetic combination model of artefenomel and piperaquine from monotherapy and combination therapy volunteer infection study data.
- **Table S7.** Parameter estimates of the pharmacokinetic/pharmacodynamic model for artefenomel and piperaquine in monotherapy and in combination.
- **Additional file 3.docx**

- **Figure S1.** Visual predictive checks for artefenomel (OZ439) concentration-time profiles from observed results when administered as monotherapy and simulations using the VIS PK model.
- **Figure S2.** Visual predictive checks for piperaquine (PQP) concentration-time profiles from observed results when administered as monotherapy and simulations using the VIS PK model.
- **Figure S3.** Visual predictive checks for artefenomel (OZ439) concentration-time profiles from observed results when administered in combination with piperaquine (PQP) and simulations using the VIS PK model.
- **Figure S4.** Visual predictive checks for piperaquine (PQP) concentration-time profiles from observed results when administered in combination with artefenomel (OZ439) and simulations using the VIS PK model.
- **Figure S5.** Individual fits for participants in the artefenomel (OZ439) + piperaquine (PQP) combination volunteer infection study (GPDI model).
- **Figure S6.** Artefenomel (OZ439) plasma concentration-time profiles by body weight of patients in the phase 2b trial compared with the PK model built from VIS data.
- **Figure S7.** Piperaquine plasma concentration-time profiles by body weight of patients in the phase 2b trial compared with the PK model built from VIS data.

