## Additional File 1 for "Characterizing the pharmacological interaction of the antimalarial combination artefenomel-piperaquine in healthy volunteers with induced blood stage *Plasmodium falciparum*"

**Text S1.** **Participant eligibility criteria**

**Inclusion criteria**

***Demography***

1. Adults (male and female) between 18 and 55 years of age inclusive, who did not live alone (from inoculation day until at least the end of the Riamet^®^ treatment) and were contactable and available for the duration of the trial and contactable up to 2 weeks following the End of Study visit (approximately 8.5 weeks).
2. Body weight minimum 50 kg, body mass index between 18 and 32 kg/m^2^, inclusive.

***Health status***

3. Certified as healthy by a comprehensive clinical assessment (detailed medical history, complete physical examination and special investigations).
4. Vital signs after 5 min resting in supine position:

- 90 mmHg ≤ systolic blood pressure ≤ 140 mmHg
- 40 mmHg ≤ diastolic blood pressure ≤ 90 mmHg
- 40 beats per min ≤ heart rate ≤ 100 beats per min.

5. Must have had QTcF ≤450 msec, QTcB ≤450 msec for male subjects, QTcF ≤470 msec, QTcB ≤470 msec for female subjects and PR interval ≤210 msec at screening and at pre-inoculation on inoculation day.
6. Heterosexual women of childbearing potential needed to be surgically sterile or using an insertable, injectable, transdermal, or combination oral contraceptive approved by the Therapeutic Goods Administration combined with a barrier contraceptive for the duration of the study, and have negative results on a urine pregnancy test done before inoculation. Abstinent, heterosexual female subjects needed to agree to start a double method if they started a sexual relationship during the study. Adequate contraception did not apply to subjects of childbearing potential with same sex partners (abstinence from penile-vaginal intercourse), when this was their preferred and usual lifestyle. Female subjects with same sex partners could not have been planning *in vitro* fertilisation within the required contraception period.

Women of non-childbearing potential who did not require contraception during the study were defined as: postmenopausal (spontaneous amenorrhoea for ≥ 12 months, or spontaneous amenorrhoea for 6–12 months and follicle-stimulating hormone (FSH) ≥ 40 IU/mL; either should be together with the absence of oral contraceptive use for > 12 months).

Male subjects participating needed to agree to use a double-barrier method of contraception including condom plus diaphragm or condom plus intrauterine device or condom plus stable oral/transdermal/injectable hormonal contraceptive by the female partner from the time of informed consent through to 90 days after the last dose of OZ439 and PQP. Abstinent male subjects needed to agree to start a double-barrier method if they began sexual relationships during the study and up to 90 days after the last dose of study drug.

Male subjects with female partners who were surgically sterile, or male subjects who had undergone sterilisation and had had testing to confirm the success of the sterilisation could also have been included.

***Regulations***

7. Having given written informed consent prior to undertaking any study-related procedure.
8. Must have been willing and able to communicate and participate in the whole study.

**Exclusion criteria**

1. Haematology, clinical chemistry, coagulation or urinalysis results at screening or on admission prior to inoculation or IMP administration that were outside of Sponsor-approved clinically acceptable laboratory ranges documented in the laboratory manual or were considered clinically relevant.
2. Any history of malaria or participation in a previous malaria challenge study.
3. Must not have travelled to or lived (>2 weeks) in a malaria-endemic region during the past 12 months or planned travel to a malaria-endemic region during the course of the study (for endemic regions see https://map.ox.ac.uk/country-profiles). Bali is not considered a malaria-endemic region.
4. Participation in any investigational product study within the 12 weeks preceding IMP administration.
5. Had evidence of increased cardiovascular disease risk (defined as >10%, 5-year risk for those greater than 35 years of age, as determined by the Australian Absolute Cardiovascular Disease Risk Calculator (http://www.cvdcheck.org.au/). Risk factors include sex, age, systolic blood pressure (mm/Hg), smoking status, total and HDL cholesterol (mmol/L), and reported diabetes status.
6. Symptomatic postural hypotension at screening on two consecutive readings, irrespective of the decrease in blood pressure, or asymptomatic postural hypotension defined as a decrease in systolic blood pressure ≥20 mmHg within 2–3 min when changing from supine to standing position.
7. History of splenectomy.
8. History or presence of diagnosed (by an allergist/immunologist) or treated (by a physician) food or known drug allergies (including but not limited to allergy to any of the antimalarial rescue medications to be used in the study), or history of anaphylaxis or other severe allergic reactions. Note: Individuals with seasonal allergies/hay fever, house dust mite, or allergy to animals that were untreated and asymptomatic at the time of dosing could have been enrolled in the study
9. History of convulsion (including intravenous drug or vaccine-induced episodes) Note: A medical history of a single febrile convulsion during childhood was not an exclusion criterion.
10. Presence of current or suspected serious chronic diseases such as cardiac or autoimmune disease (HIV or other immuno-deficiencies), insulin-dependent and non-insulin dependent diabetes (excluding glucose intolerance if exclusion criterion 4 was met), progressive neurological disease, severe malnutrition, acute or progressive hepatic disease, acute or progressive renal disease, porphyria, psoriasis, rheumatoid arthritis, asthma, epilepsy, or obsessive-compulsive disorder.
11. History of malignancy of any organ system (other than localised basal cell carcinoma of the skin or *in situ* cervical cancer), treated or untreated, within 5 years of screening, regardless of whether there was evidence of local recurrence or metastases.
12. Individuals with history of schizophrenia, bipolar disease, or other severe (disabling) chronic psychiatric diagnosis including depression or receiving psychiatric drugs or who had been hospitalised within the past 5 years prior to enrolment for psychiatric illness, history of suicide attempt, or confinement for danger to self or others.
13. History of serious psychiatric condition that may have affected participation in the study or preclude compliance with the protocol, including but not limited to past or present psychoses, disorders requiring lithium, a history of attempted or planned suicide, more than one previous episode of major depression, any previous single episode of major depression lasting for or requiring treatment for more than 6 months, or any episode of major depression during the 5 years preceding screening.

The Beck Depression Inventory was used for the assessment of depression at screening. In addition to the conditions listed above, subjects with a score of 20 or more on the Beck Depression Inventory and/or a response of 1, 2, or 3 for item 9 of this inventory (related to suicidal ideation) were not eligible for participation. These subjects were referred to a general practitioner or medical specialist as appropriate. Subjects with a Beck Depression Inventory score of 17 to 19 could have been enrolled at the discretion of the Investigator if they did not have a history of the psychiatric conditions mentioned in this criterion and their mental state was not considered to pose additional risk to the health of the subject or to the execution of the study and interpretation of the data gathered.

14. History of recurrent headache (e.g., tension-type, cluster or migraine) with a frequency of ≥2 episodes per month on average and/or severe enough to require medical therapy.
15. Presence of acute infectious disease or fever (e.g., sublingual temperature ≥38.5°C) within the 5 days prior to inoculation with malaria parasites.
16. Evidence of acute illness within the 4 weeks prior to screening that the Investigator deemed may compromise subject safety.
17. Significant inter-current disease of any type, in particular liver, renal, cardiac, pulmonary, neurologic, rheumatologic, or autoimmune disease by history, physical examination, and/or laboratory studies including urinalysis.
18. Individual had a clinically significant disease or any condition or disease that might have affected drug absorption, distribution or excretion (e.g., gastrectomy, diarrhoea).
19. Blood donation of any volume within 1 month before inclusion, or participation in any research study involving blood sampling (more than 450 mL/unit of blood), or blood donation to Australian Red Cross Blood Service (Blood Service) or other blood bank during the 8 weeks prior to the treatment drug dose in the study.
20. Individual unwilling to defer blood donations to the Blood Service for at least 6 months.
21. Medical requirement for intravenous immunoglobulin or blood transfusions.
22. Individual who had ever received a blood transfusion.
23. History or presence of alcohol abuse (alcohol consumption more than 40 g/4 units/4 standard drinks per day) or drug habituation, or any prior intravenous usage of an illicit substance.
24. Tobacco use of more than 5 cigarettes or equivalent per day, and unable to stop smoking for the duration of the clinical unit confinement.
25. Female who was breastfeeding.

***Interfering substances***

26. Any vaccination within the last 28 days.
27. Any corticosteroids, anti-inflammatory drugs, immunomodulators or anticoagulants. Any subject who was currently receiving or had previously received immunosuppressive therapy (including systemic steroids, adrenocorticotrophic hormone or inhaled steroids) at a dose or duration associated with hypothalamic-pituitary-adrenal axis suppression (e.g., 1 mg/kg/day prednisone, chronic use of inhaled high potency corticosteroids such as budesonide 800 μg/day or fluticasone 750 μg, or equivalent).
28. Any recent (<6 weeks) or current systemic therapy with an antibiotic or drug with potential antimalarial activity (e.g., chloroquine, PQP, benzodiazepine, flunarizine, fluoxetine, tetracycline, azithromycin, clindamycin, doxycycline etc.).
29. Ingestion of any poppy seeds within the 24 h prior to the screening blood test (subjects were advised by phone not to consume any poppy seeds in this time period).
30. Excessive consumption of beverages or food containing xanthine bases including Red Bull, chocolate, coffee etc. (more than 400 mg caffeine per day, equivalent to more than 4 cups of coffee per day).
31. Unwillingness to abstain from consumption of grapefruit or Seville oranges from inoculation day until end of the study.
32. Unwillingness to abstain from consumption of quinine containing foods/beverages such as tonic water and lemon bitter, from inoculation day until end of Riamet^®^ treatment.
33. Use of prescription drugs or non-prescription drugs or herbal supplements (such as St John’s Wort), within 14 days or 5 half-lives (whichever is longer) prior to the malaria inoculation. As an exception, ibuprofen (preferred) may have been used at doses of up to 1.2 g/day or paracetamol at doses of up to 4 g/day after discussion with the Investigator. Limited use of other non-prescription medications or dietary supplements, not believed to affect subject safety or the overall results of the study, may have been permitted on a case-by-case basis following approval by the Sponsor in consultation with the Investigator. Subjects were requested to refrain from taking non-approved concomitant medications from recruitment until the conclusion of the study.

***General conditions***

34. Any subject who, in the judgement of the Investigator, was likely to be non-compliant during the study, or was unable to cooperate because of a language problem or poor mental development.
35. Any subject in the exclusion period of a previous study according to applicable regulations.
36. Any subject who was the Principal Investigator or any sub-Investigator, research assistant, pharmacist, study coordinator, or other staff thereof, directly involved in conducting the study.
37. Any subject without a good peripheral venous access.

***Biological status***

38. Positive result on any of the following tests: hepatitis B surface antigen (HBs Ag), anti‑hepatitis B core antibodies (anti-HBc Ab), anti-hepatitis C virus (anti-HCV) antibodies, anti-human immunodeficiency virus 1 and 2 antibodies (anti-HIV1 and anti‑HIV2 Ab).
39. Positive urine drug test. Any drug listed in Section 7.2.1 in the urine drug screen (of the protocol) unless there is an explanation acceptable to the Investigator (e.g., the subject had stated in advance that they consumed a prescription or over-the-counter product which contained the detected drug) and/or the subject had a negative urine drug screen on retest by the pathology laboratory. Any subject who tested positive for acetaminophen (paracetamol) at screening may have still been eligible for study participation, at the Investigator’s discretion.
40. Positive alcohol breath test.

***Specific to the study***

41. Cardiac/QT risk:

- Family history of sudden death or of congenital prolongation of the QTc interval or known congenital prolongation of the QTc interval or any clinical condition known to prolong the QTc interval.
- History of symptomatic cardiac arrhythmias or with clinically relevant bradycardia.
- Electrolyte disturbances, particularly hypokalaemia, hypocalcaemia, or hypomagnesaemia.
- ECG abnormalities in the standard 12-lead ECG (at screening or at pre-inoculation on inoculation day), which in the opinion of the Investigator was clinically relevant or would interfere with the ECG analyses.

42. Known hypersensitivity to artesunate or any of its excipients, artemether or other artemisinin derivatives, PQP, proguanil/atovaquone, primaquine, or 4‑aminoquinolines.

**Text S2. Definition of adequate parasitological response on day 28 (APR_28_)**

APR_28_ was defined as a decrease in parasitemia below 1 parasite in the body before the end of day 28 (possible to evaluate only in the PK/PD model simulation) or the absence of parasitemia above the lower limit of quantification (LLOQ) on day 28, in real or PK/PD model simulated patients, whose parasitemia did not meet any of the criteria for early treatment failure (ETF) or late clinical failure (LCF) as defined hereafter. In PK/PD model simulations, the threshold parasitemia (in parasites per mL units) equivalent to 1 parasite in the body was defined by the formula:

$P_{1 parasite in the body}=\left\{ \begin{aligned} \frac{1}{W_{0}*80}, if W_{0}\leq35 kg \\ \frac{1}{W_{0}*70}, if W_{0}>35 kg \end{aligned} \right.$,

where $W_{0}$ denotes the patient weight at the beginning of the treatment.

ETF was defined as parasitemia above baseline at 48 hours or above 25% of baseline at 72 hours after start of treatment. LCF was defined as parasitemia above the LLOQ after 96 hours of treatment. For the purpose of fair comparison, the same LLOQ value of 10000 parasites per mL was assumed for both the real patient data and the PK/PD model simulated data. We note that the above definitions of ETF and LCF are solely parasitemia based simplifications of the ETF and LCF criteria defined by the World Health Organization [reference 24].
