## Additional File 2 for "Characterizing the pharmacological interaction of the antimalarial combination artefenomel-piperaquine in healthy volunteers with induced blood stage *Plasmodium falciparum*"

**Table S1.** **Volunteer infection study (VIS) data used for pharmacokinetic/ pharmacodynamic modelling of artefenomel-piperaquine combination**

| **Study** | **Dose** | **Number of participants** |
| --- | --- | --- |
| McCarthy et al., 2016 [reference 8] | Artefenomel 100 mg  Artefenomel 200 mg  Artefenomel 500 mg | 8  8  8 |
| Pasay et al., 2016 [reference 14] | Piperaquine 480 mg  Piperaquine 640 mg  Piperaquine 960 mg | 12^a^  7  5 |
| Abd-Rahman et al. [current study] | Artefenomel 200 mg/ piperaquine 320 mg  Artefenomel 200 mg/ piperaquine 480 mg  Artefenomel 200 mg/ piperaquine 640 mg  Artefenomel 400 mg/ piperaquine 480 mg  Artefenomel 400 mg/ piperaquine 640 mg  Artefenomel 800 mg/ piperaquine 640 mg  Artefenomel 800 mg/ piperaquine 960 mg | 4  2  2  2  6  4  4 |
| TOTAL | | 72 |

^a^Nine participants initially dosed with 480 mg piperaquine received a second dose of piperaquine (960 mg).

**Table S2. Phase 2b study data used in simulations to predict APR_28_ in patients**

| **Study** | **Dose** | **Number of participants^a^** | **Age (years)^c^** | **Weight (kg)^c^** | **Baseline parasitaemia (parasites/μL)^c^** |
| --- | --- | --- | --- | --- | --- |
| Macintyre et al., 2017 [reference 13] | ART 800 mg/ PQP 640 mg^b^  ART 800 mg/ PQP 960 mg^b^  ART 800 mg/ PQP 1440 mg^b^ | 108  119  119 | 11.19 (0.5-57) | 27.45 (5.6-77.2) | 25718 (187-220240) |

^a^Number of participants with pharmacokinetic data. ^b^Nominal doses for children scaled to achieve similar exposure. ^c^Mean values for all dose groups with range.

**Table S3. Adverse events by system organ class and preferred term**

| **System organ class**  Preferred term | **Overall (N=24)** | **Artefenomel 200 mg/  piperaquine 480 mg (N=2)** | **Artefenomel 200 mg/ piperaquine 640 mg (N=2)** | **Artefenomel 400 mg/ piperaquine 480 mg (N=2)** | **Artefenomel 400 mg/ piperaquine 640 mg (N=6)** | **Artefenomel 800 mg/ piperaquine 960 mg (N=4)** | **Artefenomel 200 mg/ piperaquine 320 mg (N=4)** | **Artefenomel 800 mg/ piperaquine 640 mg (N=4)** |
| --- | --- | --- | --- | --- | --- | --- | --- | --- |
|  | **Number of participants with adverse event (%) Number of adverse events** | | | | | | | |
| Any adverse event | 21 (87.5) 101 | 2 (100) 10 | 2 (100) 3 | 2 (100) 14 | 4 (66.7) 16 | 4 (100) 22 | 3 (75.0) 19 | 4 (100) 17 |
| **Nervous system disorders** | **17 (70.8) 19** | **2 (100%) 2** | **0** | **2 (100) 2** | **3 (50.0) 4** | **4 (100) 5** | **3 (75.0) 3** | **3 (75.0) 3** |
| Headache | 15 (62.5) 15 | 1 (50.0) 1 | 0 | 2 (100) 2 | 3 (50.0) 3 | 4 (100) 4 | 3 (75.0) 3 | 2 (50.0) 2 |
| Dizziness | 2 (8.3) 2 | 1 (50.0) 1 | 0 | 0 | 0 | 0 | 0 | 1 (25.0) 1 |
| Hypoaesthesia | 1 (4.2) 1 | 0 | 0 | 0 | 0 | 1 (25.0) 1 | 0 | 0 |
| Lethargy | 1 (4.2) 1 | 0 | 0 | 0 | 1 (16.7) 1 | 0 | 0 | 0 |
| **General disorders and administration site conditions** | **16 (66.7) 25** | **2 (100) 5** | **2 (100) 2** | **1 (50.0) 4** | **3 (50.0) 5** | **2 (50.0) 2** | **3 (75.0) 4** | **3 (75.0) 3** |
| Fatigue | 8 (33.3) 8 | 1 (50.0) 1 | 1 (50.0) 1 | 1 (50.0) 1 | 1 (16.7) 1 | 0 | 1 (25.0) 1 | 3 (75.0) 3 |
| Pyrexia | 7 (29.2) 7 | 2 (100) 2 | 0 | 1 (50.0) 1 | 2 (33.3) 2 | 0 | 2 (50.0) 2 | 0 |
| Vessel puncture site bruise | 5 (20.8) 5 | 0 | 1 (50.0) 1 | 1 (50.0) 1 | 1 (16.7) 1 | 2 (50.0) 2 | 0 | 0 |
| Chills | 3 (12.5) 3 | 1 (50.0) 1 | 0 | 0 | 1 (16.7) 1 | 0 | 1 (25.0) 1 | 0 |
| Feeling hot | 1 (4.2) 1 | 1 (50.0) 1 | 0 | 0 | 0 | 0 | 0 | 0 |
| Malaise | 1 (4.2) 1 | 0 | 0 | 1 (50.0) 1 | 0 | 0 | 0 | 0 |
| **Musculoskeletal and connective tissue disorders** | **12 (50.0) 17** | **1 (50.0) 1** | **1 (50.0) 1** | **2 (100) 3** | **2 (33.3) 3** | **2 (50.0) 3** | **2 (50.0) 4** | **2 (50.0) 2** |
| Myalgia | 8 (33.3) 8 | 1 (50.0) 1 | 0 | 2 (100) 2 | 1 (16.7) 1 | 1 (25.0) 1 | 2 (50.0) 2 | 1 (25.0) 1 |
| Back pain | 5 (20.8) 5 | 0 | 0 | 0 | 1 (16.7) 1 | 2 (50.0) 2 | 1 (25.0) 1 | 1 (25.0) 1 |
| Arthralgia | 2 (8.3) 2 | 0 | 0 | 1 (50.0) 1 | 1 (16.7) 1 | 0 | 0 | 0 |
| Neck pain | 1 (4.2) 1 | 0 | 0 | 0 | 0 | 0 | 1 (25.0) 1 | 0 |
| Pain in extremity | 1 (4.2) 1 | 0 | 1 (50.0) 1 | 0 | 0 | 0 | 0 | 0 |
| **Gastrointestinal disorders** | **10 (41.7) 11** | **1 (50.0) 1** | **0** | **2 (100) 3** | **0** | **4 (100) 4** | **1 (25.0) 1** | **2 (50.0) 2** |
| Nausea | 10 (41.7) 10 | 1 (50.0) 1 | 0 | 2 (100) 2 | 0 | 4 (100) 4 | 1 (25.0) 1 | 2 (50.0) 2 |
| Abdominal discomfort | 1 (4.2) 1 | 0 | 0 | 1 (50.0) 1 | 0 | 0 | 0 | 0 |
| **Investigations** | **9 (37.5) 12** | **0** | **0** | **2 (100) 2** | **1 (16.7) 1** | **1 (25.0) 1** | **3 (75.0) 5** | **2 (50.0) 3** |
| Fall in haemoglobin | 4 (16.7) 4 | 0 | 0 | 1 (50.0) 1 | 0 | 0 | 1 (25.0) 1 | 2 (50.0) 2 |
| Fall in white blood cell count | 1 (4.2) 1 | 0 | 0 | 0 | 0 | 0 | 1 (25.0) 1 | 0 |
| Fall in lymphocyte count | 4 (16.7) 4 | 0 | 0 | 1 (50.0) 1 | 0 | 0 | 2 (50.0) 2 | 1 (25.0) 1 |
| Fall in neutrophil count | 2 (8.3) 2 | 0 | 0 | 0 | 1 (16.7) 1 | 0 | 1 (25.0) 1 | 0 |
| Prolongation of QT interval | 1 (4.2) 1 | 0 | 0 | 0 | 0 | 1 (25.0) 1 | 0 | 0 |
| **Injury, poisoning and procedural complications** | **5 (20.8) 5** | **0** | **0** | **0** | **1 (16.7) 1** | **3 (75.0) 3** | **1 (25.0) 1** | **0** |
| Contusion | 2 (8.3%) 2 | 0 | 0 | 0 | 0 | 2 (50.0) 2 | 0 | 0 |
| Sunburn | 1 (4.2) 1 | 0 | 0 | 0 | 0 | 1 (25.0) 1 | 0 | 0 |
| Vascular access site bruising | 1 (4.2) 1 | 0 | 0 | 0 | 1 (16.7) 1 | 0 | 0 | 0 |
| Wrist fracture | 1 (4.2) 1 | 0 | 0 | 0 | 0 | 0 | 1 (25.0) 1 | 0 |
| **Infections and infestations** | **4 (16.7) 4** | **0** | **0** | **0** | **0** | **1 (25.0) 1** | **1 (25.0) 1** | **2 (50.0) 2** |
| Upper respiratory tract infection | 2 (8.3) 2 | 0 | 0 | 0 | 0 | 0 | 0 | 2 (50.0) 2 |
| Rhinovirus infection | 1 (4.2) 1 | 0 | 0 | 0 | 0 | 0 | 1 (25.0) 1 | 0 |
| Viral infection | 1 (4.2) 1 | 0 | 0 | 0 | 0 | 1 (25.0) 1 | 0 | 0 |
| **Cardiac disorders** | **2 (8.3) 2** | **1 (50.0) 1** | **0** | **0** | **0** | **0** | **0** | **1 (25.0) 1** |
| Tachycardia | 2 (8.3) 2 | 1 (50.0) 1 | 0 | 0 | 0 | 0 | 0 | 1 (25.0) 1 |
| Metabolism and nutrition disorders | 2 (8.3) 2 | 0 | 0 | 0 | 0 | 1 (25.0) 1 | 0 | 1 (25.0) 1 |
| Decreased appetite | 2 (8.3%) 2 | 0 | 0 | 0 | 0 | 1 (25.0) 1 | 0 | 1 (25.0) 1 |
| **Respiratory, thoracic and mediastinal disorders** | **2 (8.3) 2** | **0** | **0** | **0** | **1 (16.7) 1** | **1 (25.0) 1** | **0** | **0** |
| Dry throat | 1 (4.2) 1 | 0 | 0 | 0 | 0 | 1 (25.0) 1 | 0 | 0 |
| Nasal congestion | 1 (4.2) 1 | 0 | 0 | 0 | 1 (16.7) 1 | 0 | 0 | 0 |
| **Eye disorders** | **1 (4.2) 1** | **0** | **0** | **0** | **0** | **1 (25.0) 1** | **0** | **0** |
| Dry eye | 1 (4.2) 1 | 0 | 0 | 0 | 0 | 1 (25.0) 1 | 0 | 0 |
| **Skin and subcutaneous tissue disorders** | **1 (4.2) 1** | **0** | **0** | **0** | **1 (16.7) 1** | **0** | **0** | **0** |
| Contact dermatitis | 1 (4.2) 1 | 0 | 0 | 0 | 1 (16.7) 1 | 0 | 0 | 0 |

Adverse events were coded to system organ class and preferred term using MedDRA^®^ Version 21.0.

**Table S4. Plasma artefenomel and piperaquine non-compartmental pharmacokinetic parameters**

| **Parameter** | **Artefenomel 200 mg/ piperaquine 480 mg**  **[N=2]** | **Artefenomel 200 mg/ piperaquine 640 mg**  **[N=2]** | **Artefenomel 400 mg/ piperaquine 480 mg**  **[N=2]** | **Artefenomel 400 mg/ piperaquine 640 mg**  **[N=6]** | **Artefenomel 800 mg/ piperaquine 960 mg**  **[N=4]** | **Artefenomel 200 mg/ piperaquine 320 mg**  **[N=4]** | **Artefenomel 800 mg/ piperaquine 640 mg**  **[N=4]** |
| --- | --- | --- | --- | --- | --- | --- | --- |
| **Artefenomel** | | | | | | | |
| C_max_ (ng/L) | 0.4145 (41.4) | 0.2582 (22.9) | 0.8829 (48.2) | 0.6835 (32.7) | 2.135 (38.0) | 0.3208 (113.6) | 1.910 (15.3) |
| t_max_ (h) | 2.5 (2.0,3.0) | 2.5 (2.0,3.0) | 3.0 (3.0,3.0) | 2.5 (2.0,3.0) | 2.5 (2.0,3.0) | 2.0 (2.0,6.0) | 3.0 (3.0,3.0) |
| AUC_0-last_ (h*ng/L) | 2.861 (91.4) | 1.763 (27.0) | 8.734 (60.7) | 6.245 (25.5) | 20.29 (32.4) | 1.582 (72.7) | 19.39 (6.8) |
| AUC_0-inf_ (h*ng/L) | NR | NR | 9.056 (57.3) | 6.702 (29.9) | 23.84 (13.1)^b^ | NR | 19.78 (6.6) |
| t_1/2_ (h) | NR | NR | 146.4 (2.9) | 147.1 (68.8) | 161.5 (25.8)^b^ | NR | 168.4 (49.0) |
| CL/F (L/h) | NR | NR | 44.17 (57.3) | 59.68 (29.9) | 33.55 (13.1)^b^ | NR | 40.44 (6.6) |
| Vz/F (L) | NR | NR | 9330 (53.7) | 12660 (39.8) | 7818 (33.0)^b^ | NR | 9825 (52.3) |
| **Piperaquine** | | | | | | | |
| C_max_ (ng/L) | 0.05441 (39.0) | 0.1168 (31.3) | 0.06458 (45.9) | 0.1048 (99.6) | 0.4104 (19.7) | 0.04609 (124.2) | 0.1860 (100.1) |
| t_max_ (h) | 4.0 (3.0,5.0) | 5.0 (4.0,6.0) | 3.5 (3.0,4.0) | 4.0 (2.0, 16.0) | 3.0 (2.0,4.0) | 4.0 (2.0,12.2) | 4.5 (2.0,24.0) |
| AUC_0-last_ (h*ng/L) | 3.778 (0.5) | 8.760 (15.8) | 3.575 (50.5) | 5.794 (30.0) | 16.14 (6.1) | 2.386 (60.6) | 10.49 (43.8) |
| AUC_0-inf_ (h*ng/L) | 4.257 (3.3) | NR | NR | 8.169 (ND)^a^ | 20.25 (0.7)^c^ | 3.803 (17.0)^c^ | 16.70 (17.1)^c^ |
| t_1/2_ (h) | 262.3 (12.7) | NR | NR | 393.6 (ND)^a^ | 395.9 (6.8)^c^ | 267.2 (25.8)^c^ | 253.2 (8.9)^c^ |
| CL/F (L/h) | 112.8 (3.3) | NR | NR | 78.35 ND)^a^ | 47.42 (0.7)^c^ | 84.15 (17.0)^c^ | 38.32 (17.1)^c^ |
| Vz/F (L) | 42670 (9.4) | NR | NR | 44480 (ND)^a^ | 27080 (6.1)^c^ | 32430 (8.6)^c^ | 14000 (26.3)^c^ |

Data are geometric means (coefficient of variation [%]) except t_max_ which is median (minimum, maximum). C_max_: maximum observed concentration; t_max_: time to reach the maximum observed concentration; AUC_0-last_: area under the concentration-time curve from time 0 (dosing) to the last sampling time at which the concentration is at or above the lower limit of quantification; AUC_0-inf_: area under the concentration-time curve from time 0 (dosing) extrapolated to infinity; t_½_: apparent terminal half-life; CL/F: apparent total clearance; Vz/F: apparent volume of distribution; NR: not reported; ND: not determined. Calculation based on n=1^a^, n=3^b^, or n=2^c^. Values for AUC_0-inf_, t_1/2_, CL/F, and Vz/F were only reported for a participant if the following criteria were met: a minimum of 3 measurable concentration-time points during the log-linear portion of the terminal elimination phase (excluding C_max_); r^2^ > 0.80 for the regression of the log concentration-time data during the terminal elimination phase; negative slope for log regression fit; extrapolated portion of AUC_0-inf_ < 20% of total AUC_0-inf_.

**Table S5. Individual participant parasite clearance parameters**

| **Participant ID** | **Log_10_PRR_48_ (95% CI)** | **PCt_1/2_ (95% CI)** | **Parasite regrowth*** |
| --- | --- | --- | --- |
| **Artefenomel 200 mg/ piperaquine 480 mg** | | |  |
| 103 | 2.3 (2.07 - 2.52) | 6.29 (5.73 - 6.97) | Yes (day 22) |
| 106 | 1.98 (1.69 - 2.27) | 7.29 (6.36 - 8.55) | Yes (day 12) |
| **Artefenomel 200 mg/ piperaquine 640 mg** | | | |
| 102 | 2.17 (1.88 - 2.45) | 6.67 (5.89 - 7.69) | No |
| 104 | 3.01 (2.44 - 3.57) | 4.8 (4.04 - 5.91) | No |
| **Artefenomel 400 mg/ piperaquine 480 mg** | | | |
| 101 | 3.28 (2.96 - 3.59) | 4.41 (4.02 - 4.88) | No |
| 108 | 4.84 (4.2 - 5.49) | 2.98 (2.63 - 3.44) | Yes (day 22) |
| **Artefenomel 400 mg/ piperaquine 640 mg** | | |  |
| 105 | 3.39 (2.85 - 3.93) | 4.26 (3.67 - 5.07) | No |
| 107 | 3.3 (2.9 - 3.7) | 4.38 (3.9 - 4.99) | No |
| 302 | 3.33 (2.96 - 3.69) | 4.34 (3.91 - 4.88) | No |
| 306 | 4.17 (3.68 - 4.66) | 3.46 (3.1 - 3.93) | No |
| 307 | 5.66 (5.03 - 6.28) | 2.55 (2.3 - 2.87) | No |
| 308 | 3.62 (3.26 - 3.98) | 3.99 (3.63 - 4.43) | No |
| **Artefenomel 800 mg/ piperaquine 960 mg** | | | |
| 201 | 2.81 (2.31 - 3.31) | 5.14 (4.37 - 6.24) | No |
| 205 | 4.32 (3.98 - 4.67) | 3.34 (3.1 - 3.63) | No |
| 206 | 4.52 (3.37 - 5.67) | 3.2 (2.55 - 4.29) | No |
| 207 | 4.5 (3.91 - 5.09) | 3.21 (2.84 - 3.7) | No |
| **Artefenomel 200 mg/ piperaquine 320 mg** | | |  |
| 202 | 3.13 (2.84 - 3.42) | 4.62 (4.22 - 5.09) | Yes (day 13) |
| 203 | 1.2 (0.96 - 1.44) | 12.04 (10.06 - 14.99) | Yes (day 13) |
| 204 | 2.19 (1.83 - 2.56) | 6.59 (5.65 - 7.89) | Yes (day 13) |
| 208 | 1.56 (1.28 - 1.84) | 9.24 (7.84 - 11.26) | Yes (day 13) |
| **Artefenomel 800 mg/ piperaquine 640 mg** | | | |
| 301 | 3.46 (3.01 - 3.92) | 4.17 (3.69 - 4.81) | No |
| 303 | 3.38 (2.99 - 3.77) | 4.28 (3.84 - 4.84) | No |
| 304 | 4.88 (4.15 - 5.61) | 2.96 (2.58 - 3.48) | No |
| 305 | 3.49 (3.2 - 3.79) | 4.14 (3.82 - 4.52) | No |

Log_10_PRR_48_: parasite reduction ration over a 48 hour period on a log scale; PCt_1/2_: parasite clearance half-life in hours; CI: confidence interval. *The day of parasite regrowth is defined as the day prior to the day on which artemether-lumefantrine treatment was initiated (day relative to inoculation with the malaria challenge agent).

**Table S6. Parameter estimates for the final pharmacokinetic combination model of artefenomel and piperaquine from monotherapy and combination therapy volunteer infection study data**

| PARAMETER | VALUE | RSE | SHRINKAGE | COMMENT |
| --- | --- | --- | --- | --- |
| **Typical parameters** |  |  |  |  |
| Fabs1x1 | 1 (FIX) | - | - | Relative bioavailability artefenomel (-) |
| kax1 | 0.216 | 4.05% | - | Absorption rate parameter artefenomel (1/hour) |
| CLx1 | 68 | 5.97% | - | Apparent clearance artefenomel (L/hour) |
| Vcx1 | 81.5 | 12.2% | - | Apparent central volume artefenomel (L) |
| Q1x1 | 10.6 | 4.32% | - | Apparent intercompartmental clearance artefenomel (L/hour) |
| Vp1x1 | 1070 | 7.61% | - | Apparent peripheral volume artefenomel (L) |
| Fabs1x2 | 1 (FIX) | - | - | Relative bioavailability piperaquine (-) |
| kax2 | 0.0794 | 9.32% | - | Absorption rate parameter piperaquine (1/hour) |
| CLx2 | 72.3 | 7.28% | - | Apparent clearance piperaquine (L/hour) |
| Vcx2 | 582 | 11.8% | - | Apparent central volume piperaquine (L) |
| Q1x2 | 334 | 7.38% | - | Apparent intercompartmental clearance piperaquine (L/hour) |
| Vp1x2 | 30800 | 8.11% | - | Apparent peripheral volume piperaquine (L) |
| Tlag1 | 0.409 | 2.18% | - | - |
| Tlag2 | 0.39 | 2.8% | - | - |
| **Inter-individual variability** |  |  |  |  |
| omega(Fabs1x1) | 0 (FIX) | - | - | Normal |
| omega(kax1) | 0.0897 | 42.2% | 61% | LogNormal |
| omega(CLx1) | 0.348 | 11.3% | 21% | LogNormal |
| omega(Vcx1) | 0.687 | 12.8% | 29% | LogNormal |
| omega(Q1x1) | 0.05 (FIX) | - | 74% | LogNormal |
| omega(Vp1x1) | 0.572 | 21.9% | 24% | LogNormal |
| omega(Fabs1x2) | 0 (FIX) | - | - | Normal |
| omega(kax2) | 0.495 | 13.4% | 27% | LogNormal |
| omega(CLx2) | 0.461 | 11.9% | 22% | LogNormal |
| omega(Vcx2) | 0.463 | 23.2% | 38% | LogNormal |
| omega(Q1x2) | 0.409 | 14.1% | 31% | LogNormal |
| omega(Vp1x2) | 0.505 | 12.5% | 25% | LogNormal |
| omega(Tlag1) | 0.104 | 17.6% | 31% | LogNormal |
| omega(Tlag2) | 0.144 | 13.6% | 29% | LogNormal |
| **Correlation of random effects** |  |  |  |  |
| corr(CLx1,Vcx1) | -0.148 | 110% | - | Correlation coefficient |
| corr(CLx2,Vcx2) | 0.577 | 33.9% | - | Correlation coefficient |
| **Parameter-Covariate relationships** |  |  |  |  |
| beta_kax1(AUC1) | -0.377 | 11.6% | - | AUC of artefenomel in mg/kg^0.75 on kax1 |
| beta_CLx1(AUC1) | -0.383 | 16.7% | - | AUC of artefenomel in mg/kg^0.75 on CLx1 |
| beta_CLx1(WT0) | 0.75 (FIX) | - | - | Weight in kg on CLx1 |
| beta_Vcx1(WT0) | 1 (FIX) | - | - | Weight in kg on Vcx1 |
| beta_Q1x1(WT0) | 0.75 (FIX) | - | - | Weight in kg on Q1x1 |
| beta_Vp1x1(WT0) | 1 (FIX) | - | - | Weight in kg on Vp1x1 |
| beta_CLx2(WT0) | 0.75 (FIX) | - | - | Weight in kg on CLx2 |
| beta_Vcx2(WT0) | 1 (FIX) | - | - | Weight in kg on Vcx2 |
| beta_Q1x2(WT0) | 0.75 (FIX) | - | - | Weight in kg on Q1x2 |
| beta_Vp1x2(WT0) | 1 (FIX) | - | - | Weight in kg on Vp1x2 |
| **Residual Variability** |  |  |  |  |
| error_PROP1 | 0.354 | 2.67% | - | Proportional Error (fraction) - Concentration artefenomel (ug/mL) |
| error_PROP2 | 0.366 | 2.68% | - | Proportional Error (fraction) - Concentration piperaquine (ug/mL) |
| Objective function | -9757 | - | - | - |
| AIC | -9699 | - | - | - |
| BIC | -9633 | - | - | - |

Objective function rounded to closest integer value, omega values reported in standard deviation.

**Table S7. Parameter estimates of the pharmacokinetic/pharmacodynamic model for artefenomel and piperaquine in monotherapy and in combination**

| **Parameters** | **Description** | **Estimates^a^** |
| --- | --- | --- |
| *Natural growth* | | |
| PL_base_ (ln parasites) | Baseline parasitemia | -3.64 (19) |
| k_grow_ (/h) | Growth rate constant | 0.0709 (6.22) |
| IIV PL_base_ | Interindividual variability in PL_base_ | 2.5 (17)^b^ |
| IIV k_grow_ | Interindividual variability in k_grow_ | 0.221 (15.4)^b^ |
| *Artefenomel exposure-effect relationship monotherapy* | | |
| EMAXx1 (/h) | Maximum killing rate of artefenomel | 0.194 (10.3) |
| EC50x1 (µg/mL) | Artefenomel concentration at which half EMAXx1 is reached | 0.00289 (26.7) |
| hillx1 | Steepness of the concentration-effect relationship | 3.09 (6.35) |
| *Piperaquine exposure-effect relationship monotherapy* | | |
| EMAXx2 (/h) | Maximum killing rate of piperaquine | 0.262 (3.82) |
| EC50x2 (µg/mL) | Piperaquine concentration at which half EMAXx2 is reached | 0.00626 (17.9) |
| hillx2 | Steepness of the concentration-effect relationship | 5.24 (5.32) |
| *Pharmacodynamic interaction (GPDI model)* | | |
| INT_EC50_ | Maximal fractional change of EC_50_ | -0.117 (7.12) |
| INT_Emax_ | Maximal fractional change of E_max_ | -0.406 (8.19) |
| EC50_INT12, EC50_ (µg/mL) | Potency of the EC_50_ interaction of artefenomel mediated by piperaquine | 0.00626 (fixed) |
| EC50_INT21, EC50_ (µg/mL) | Potency of the EC_50_ interaction of piperaquine mediated by artefenomel | 0.000196 (fixed) |
| EC50_INT12, Emax_ (µg/mL) | Potency of the E_max_ interaction of artefenomel mediated by piperaquine | 0.00626 (fixed) |
| EC50_INT21, Emax_ (µg/mL) | Potency of the E_max_ interaction of piperaquine mediated by artefenomel | 0.000196 (fixed) |
| *Residual error* | | |
| Proportional error |  | 1.29 (3.4) |

^a^ Value in estimate (relative standard error) unless otherwise stated.

^b^ Value in standard deviation (relative standard error).
