## Additional File 3 for "Characterizing the pharmacological interaction of the antimalarial combination artefenomel-piperaquine in healthy volunteers with induced blood stage *Plasmodium falciparum*"

**Figure S1. Visual predictive checks for artefenomel (OZ439) concentration-time profiles from observed results when administered as monotherapy and simulations using the VIS PK model**

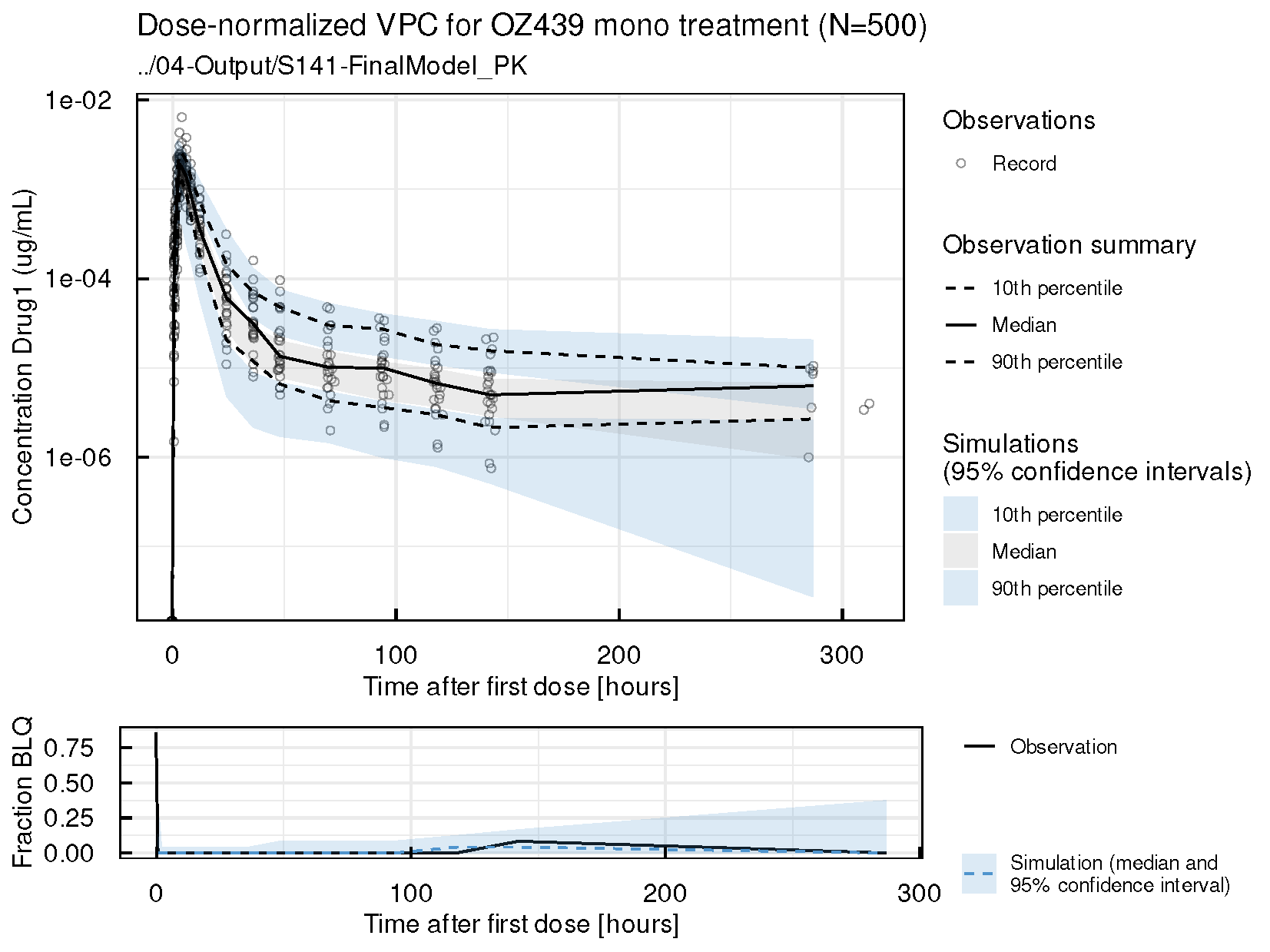

Drug 1= Artefenomel (OZ439)

**Figure S2. Visual predictive checks for piperaquine (PQP) concentration-time profiles from observed results when administered as monotherapy and simulations using the VIS PK model**

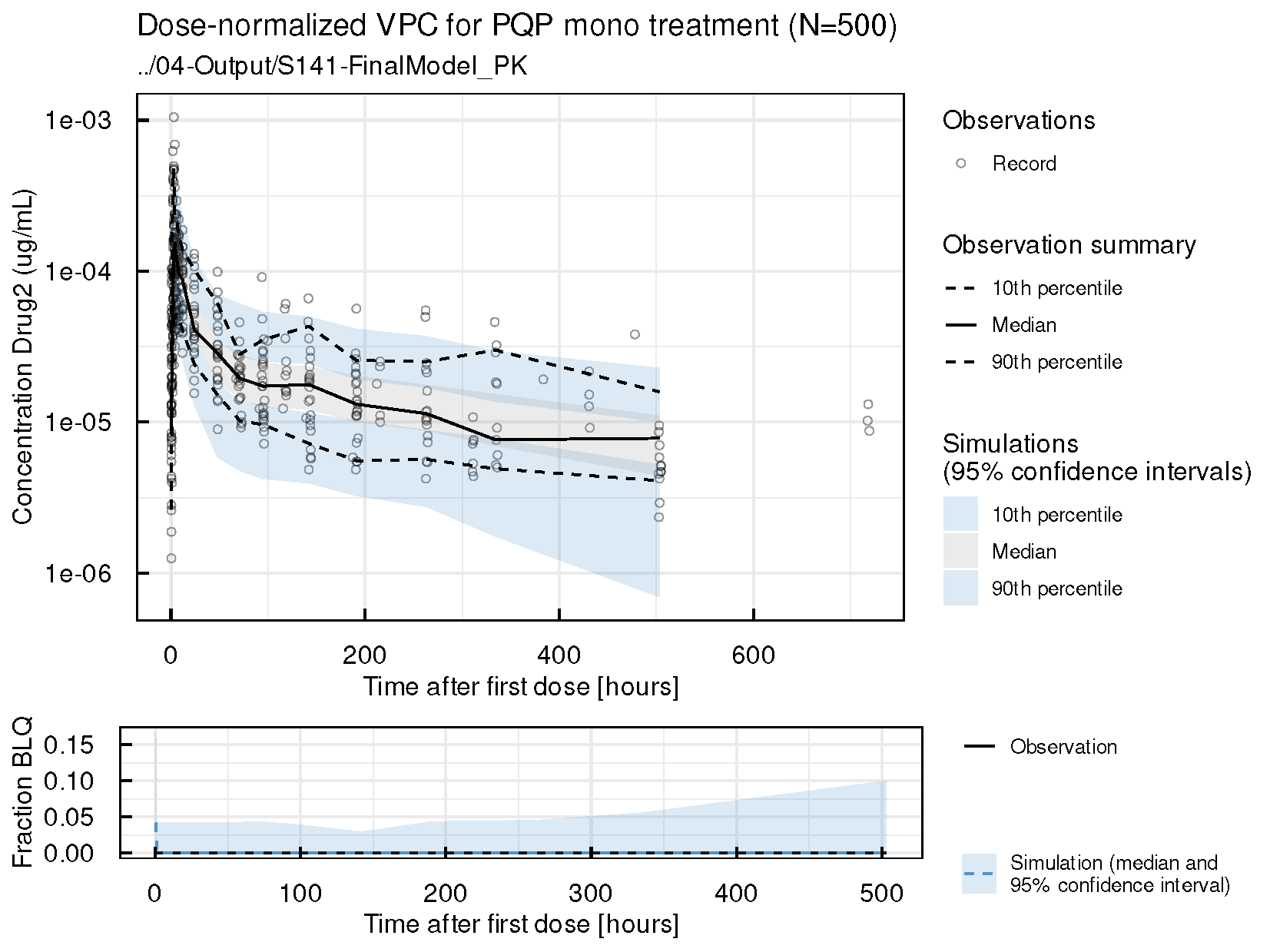

Drug 2= Piperaquine

**Figure S3. Visual predictive checks for artefenomel (OZ439) concentration-time profiles from observed results when administered in combination with piperaquine (PQP) and simulations using the VIS PK model**

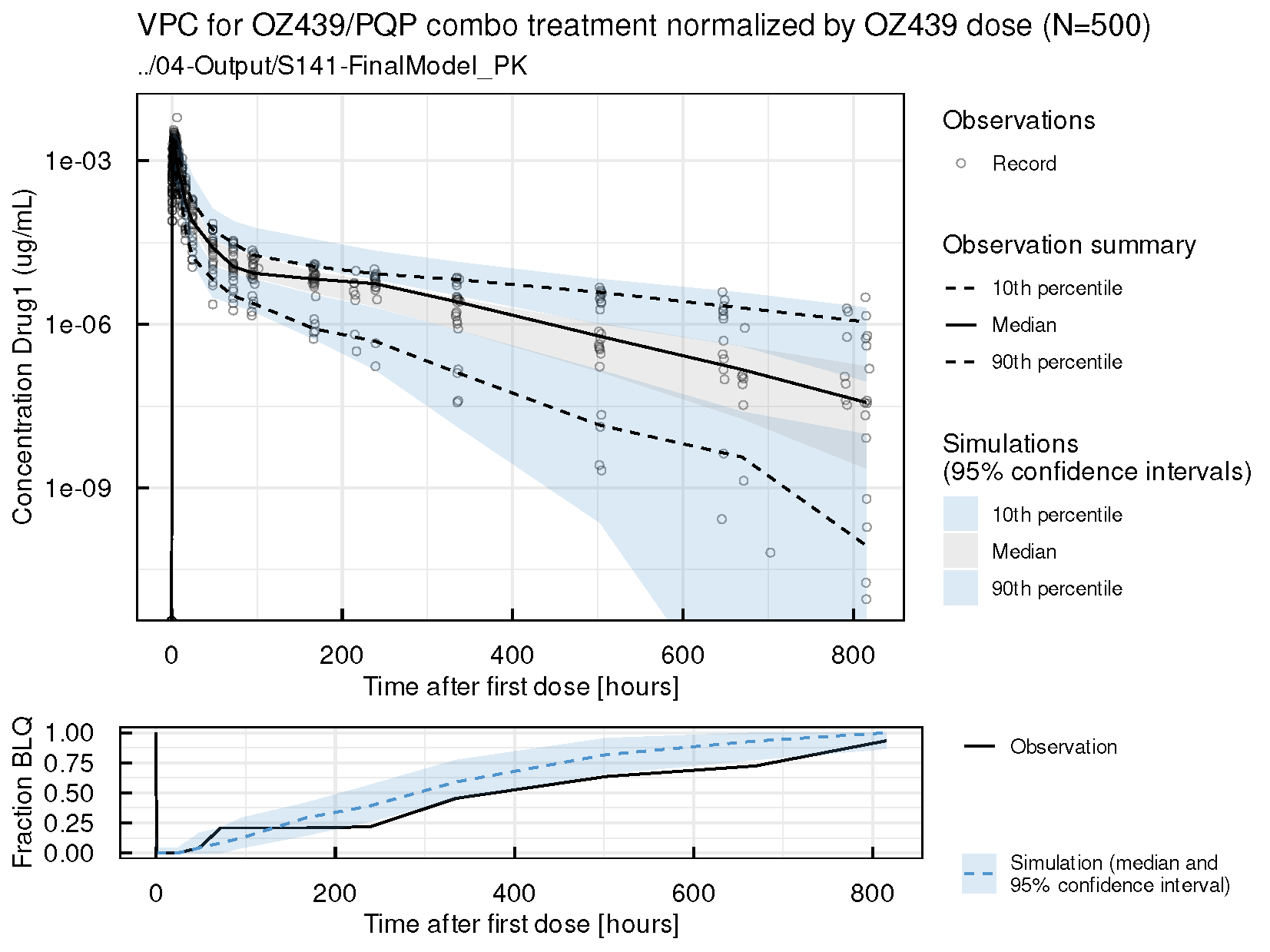

Drug 1= Artefenomel (OZ439)

**Figure S4. Visual predictive checks for piperaquine (PQP) concentration-time profiles from observed results when administered in combination with artefenomel (OZ439) and simulations using the VIS PK model**

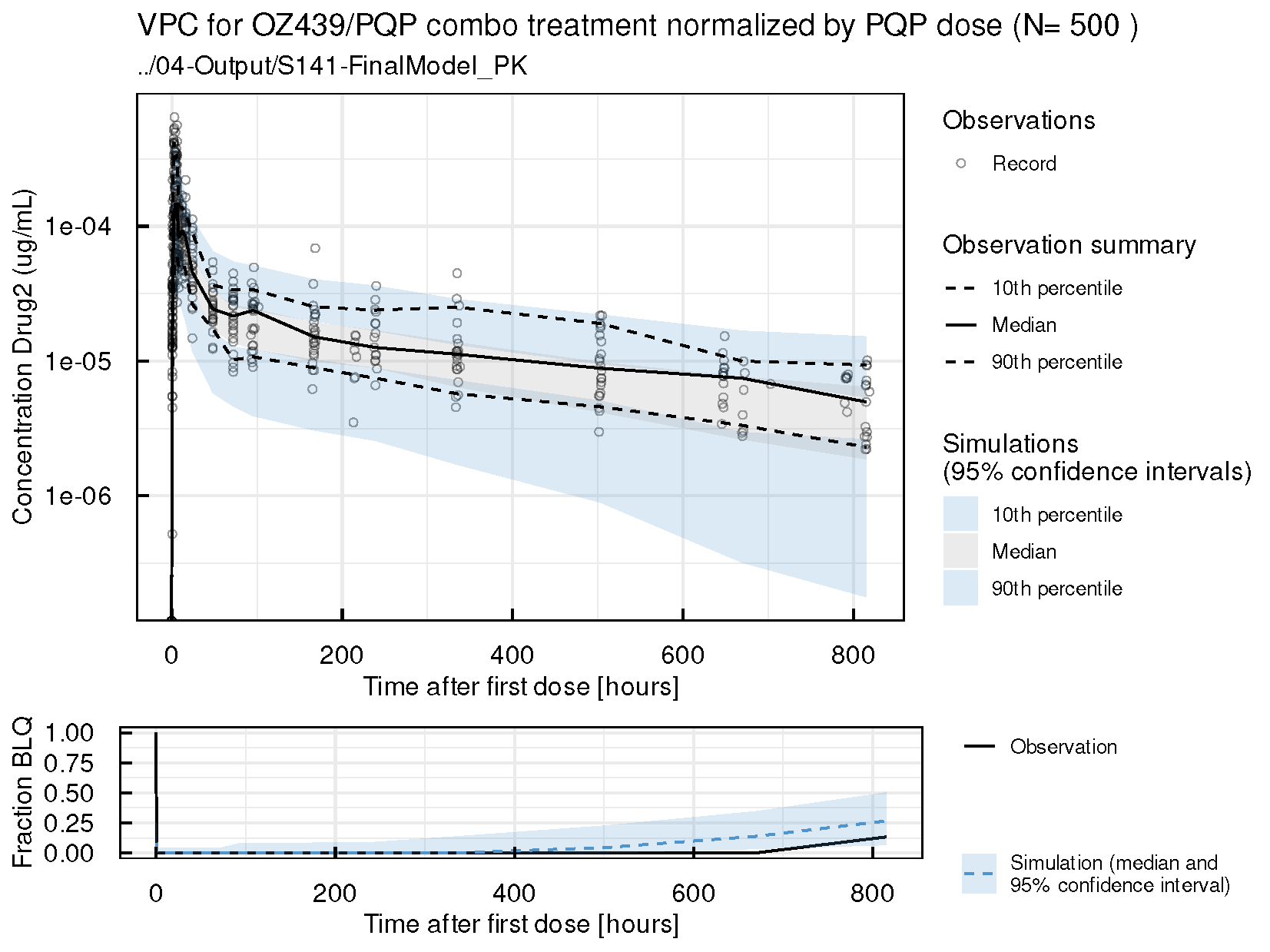

Drug 2= Piperaquine

**Figure S5. Individual fits for participants in the artefenomel (OZ439) + piperaquine (PQP) combination volunteer infection study (GPDI model)**

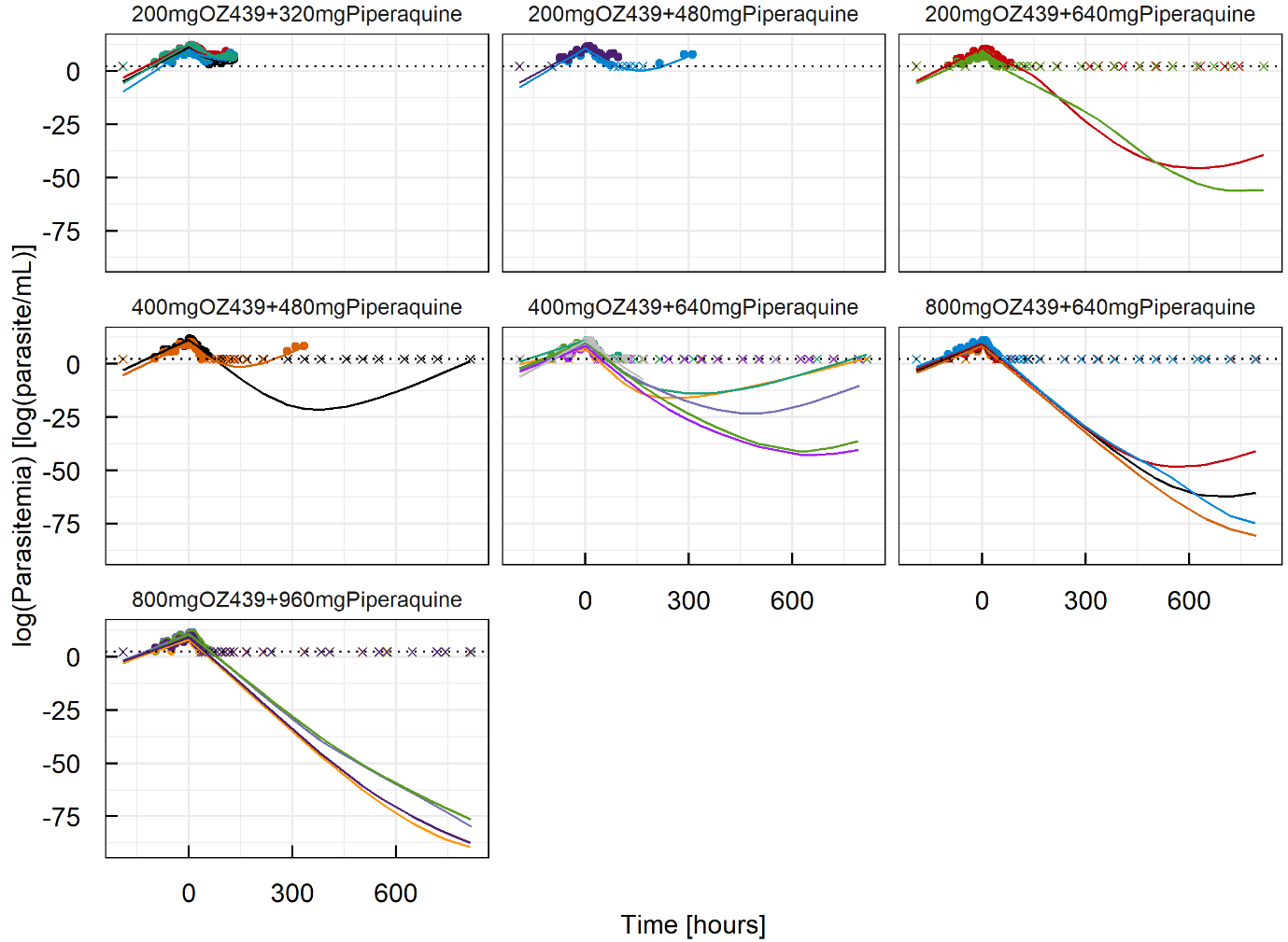

Lines represent individual predictions, dots represent observed parasitaemia values, and crosses represent observed parasitaemia below the lower limit of quantification. Colours indicate different participants.

**Figure S6. Artefenomel (OZ439) plasma concentration-time profiles by body weight of patients in the phase 2b trial compared with the PK model built from VIS data**

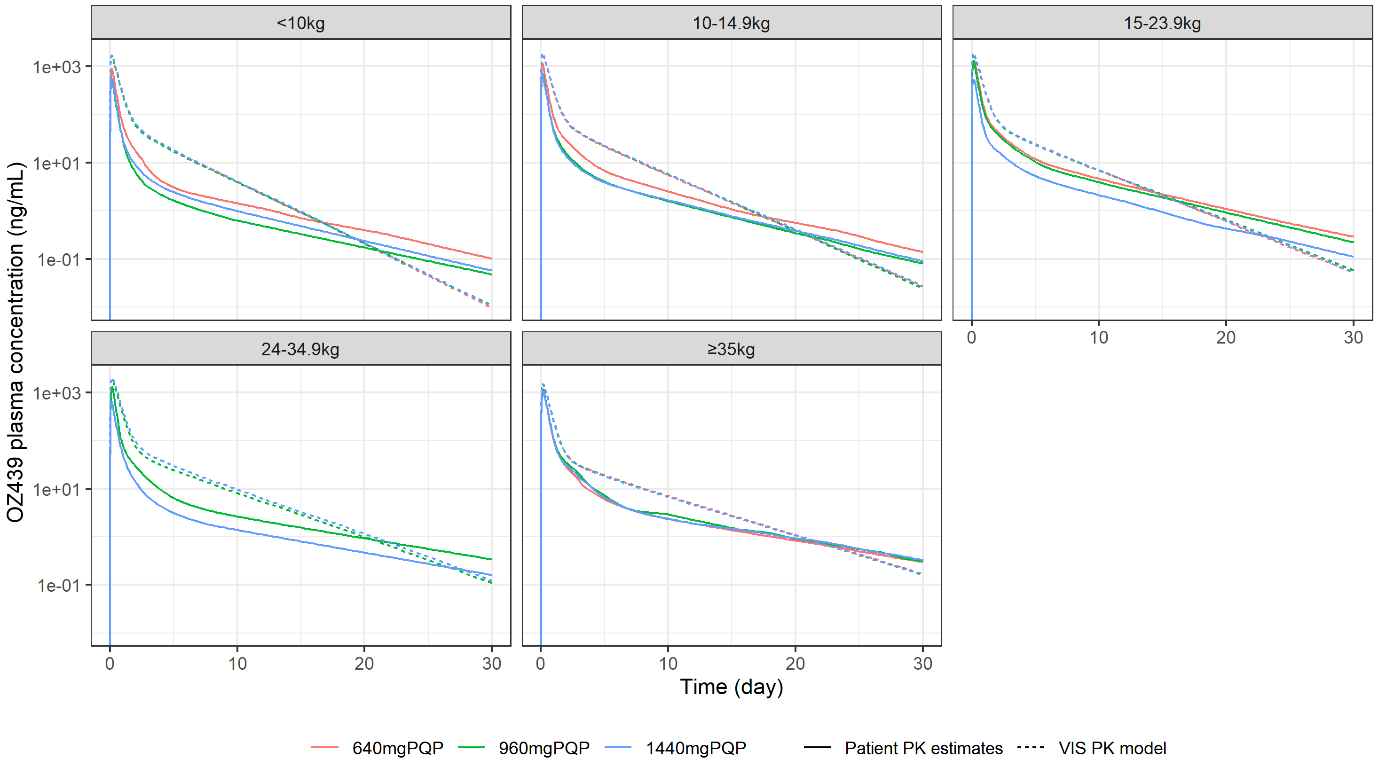

**Figure S7. Piperaquine plasma concentration-time profiles by body weight of patients in the phase 2b trial compared with the PK model built from VIS data**

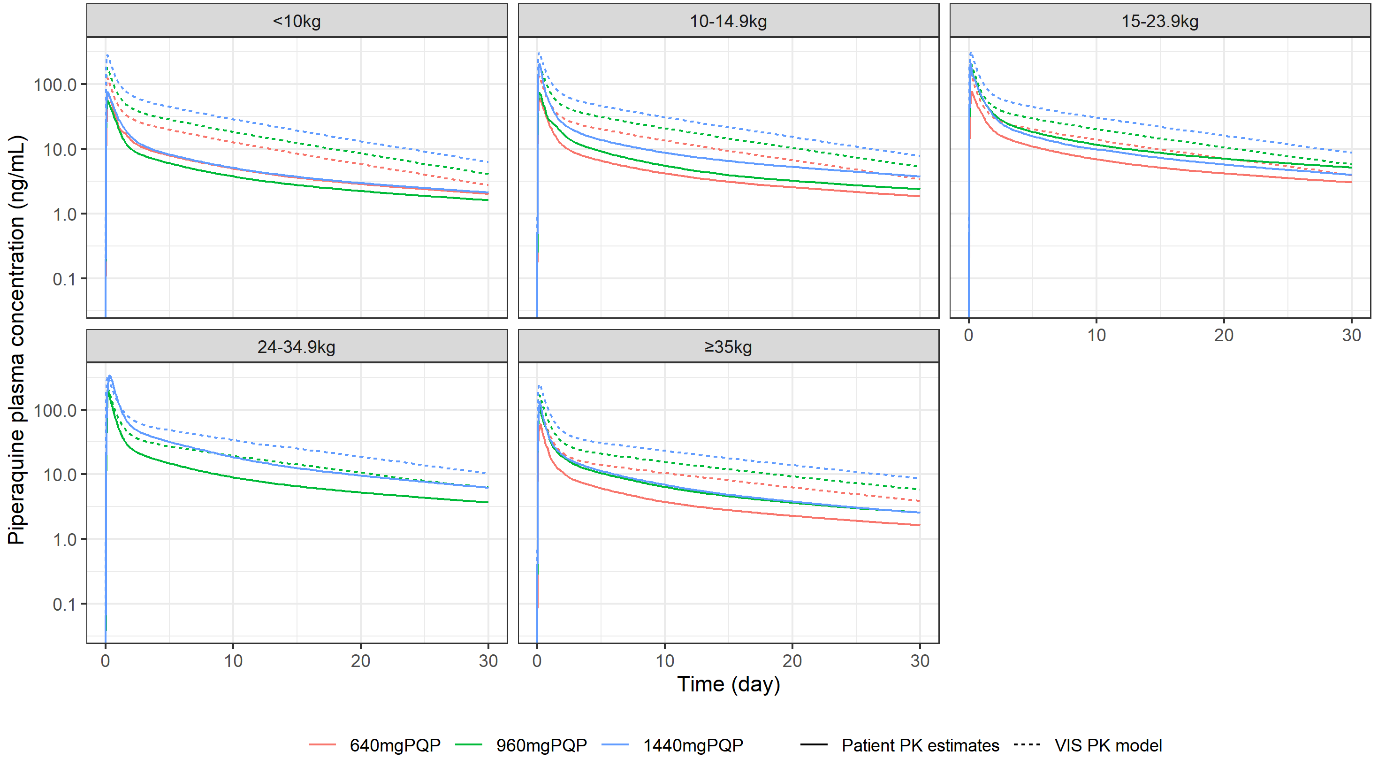
